# Sanitation-related empowerment resources are associated with women’s well-being, anxiety, and depression: findings from Bangladesh, India, Senegal, and Uganda

**DOI:** 10.1101/2025.01.08.25320223

**Authors:** Thea Mink, Madeleine Patrick, Amelia Conrad, Tanvir Ahmed, Srishty Arun, Vinod Ramanarayanan, Niladri Chakraborti, Y. Malini Reddy, Abhilaasha Nagarajan, Tanushree Bhan, Sheela S. Sinharoy, Bethany A. Caruso

**Author notes:** Corresponding author, (BAC). Authors contributed equally (Sheela S. Sinharoy, Bethany A. Caruso).

## Abstract

Nascent public health research has identified linkages between sanitation experiences and mental health. The present study examined associations between sanitation-related empowerment resources (Bodily Integrity, Safety and Security, Privacy, and Time) and mental health outcomes (well-being, depression, and anxiety).

We conducted a secondary analysis of cross-sectional data collected in 2021-2022 from household surveys of women in Bangladesh, India, Senegal, and Uganda (n = 2,122). The primary exposures were sanitation-related empowerment resources measured using the Agency, Resources, and Institutional Structures for Sanitation-related Empowerment (ARISE) Scales. Three outcomes were assessed: subjective well-being (World Health Organization Well-being Index, WHO-5), anxiety (General Anxiety Disorder measure, GAD-2), and depression (Patient Health Questionnaire, PHQ-2). Linear regressions of the WHO-5, PHQ-2, and GAD-2 scores on the four sanitation-related empowerment resources were conducted.

Overall mean scores for well-being were moderate, and overall mean scores for anxiety and depression indicated normal prevalence (well-being mean = 17.2, SD = 5.8, depression mean = 1.1, SD = 1.4; anxiety mean = 1.0, SD = 1.4). In the adjusted well-being model, there was a positive association between Privacy and well-being (β = 2.0, p<.001). In the adjusted depression model, there were negative associations between Bodily Integrity and depression (β = -0.3, p=.002) and between Privacy and depression (β = -0.4, p<.001). In the adjusted anxiety model, there were negative associations between all four resources and anxiety (Bodily Integrity β = -0.7, p<.001; Safety and Security β = -0.3, p=.025; Privacy β = - 0.3, p=.037; Time β = -0.2, p=.009).

Our findings provide evidence of associations between women’s sanitation-related resources of Bodily Integrity, Safety and Security, Privacy, and Time and mental health. Sanitation initiatives should aim to enhance and evaluate women’s experiences of these resources given their potential to benefit women’s mental health and well-being.

## 1 Introduction

Sanitation is a human right and is considered essential for preventing infectious diseases and contributing to mental and social well-being [1, 2]. Despite the critical role of sanitation for human health, 3.5 billion people still lack access to safely managed sanitation, including an estimated 419 million who practice open defecation [3]. While the World Health Organization (WHO) acknowledges the importance of sanitation on mental and social well-being, most research to date has focused on infectious disease outcomes, including understanding the burden of poor sanitation on various infectious diseases and related sequelae, like stunting, and determining what interventions are most effective at preventing diarrheal disease [4, 5]. There is also a growing understanding that inadequate sanitation can pose broader risks to women’s health in particular, including maternal mortality, gender-based violence and harassment, and unmet menstrual health needs [6–11]. As a result, there has been a call for more research on non-infectious disease outcomes, including mental health and general well-being, to more comprehensively understand how sanitation can impact health [12].

A small but growing body of research has identified linkages between negative sanitation experiences and adverse mental health outcomes. A systematic review and qualitative synthesis investigating how sanitation influences mental health found that an absence of sanitation-related privacy and safety negatively influenced well-being [13]. Specifically, women reported feeling anxiety when they perceived that they were exposed or were at risk of verbal, physical, or sexual assault. Qualitative studies have documented that sanitation-related conditions, like the physical environment (e.g., barriers to access), social restrictions and conflict, and sexual violence are related to women’s psychosocial stress [14–16]. Additionally, a quantitative study conducted with women in rural Odisha, India found women’s negative experiences and concerns, or ‘sanitation insecurity,’ to be associated with depression, anxiety, and poor well-being [17]. Notably, these associations were present even when women had access to a functional household latrine. This assessment was carried out using a measure of ‘sanitation insecurity’ that was created for the population engaged in rural Odisha, limiting comparability across other settings. Taken together, these findings provide an important foundation for understanding how women’s sanitation experiences can influence mental health. However, further research is needed, particularly quantitative research that investigates sanitation experiences across diverse populations.

Research and practice are also increasingly seeking to understand the connection between sanitation and women’s empowerment [18]. Empowerment is widely acknowledged to be a complex and multi-dimensional construct, and the ‘resources’ dimension of empowerment may be most relevant to research on sanitation and mental health [18]. In the women’s empowerment literature, Kabeer defined resources as, “not only access, but also future claims, to both material and human and social resources” [19]. Similarly, the KIT Institute’s (KIT) conceptual framework of women’s empowerment defines resources as the, “tangible and intangible capital and sources of power that women and girls have, own or use individually or collectively in the exercise of agency” [20]. The framework includes three main types of resources that are central to women’s and girls’ empowerment: Critical Consciousness, Assets (includes Social Capital, Knowledge and Skills, Time, Financial and Productive Assets), and Bodily Integrity (includes Health, Safety and Security). A recent systematic review leveraged and adapted the KIT framework to understand how sanitation research has engaged empowerment and related domains and sub-domains [18]. From the review, the resources sub-domains of Bodily Integrity, Safety and Security, Privacy, and Time were the most amenable to programmatic change and seemed to have the most potential to be linked to mental health outcomes [18].

The literature suggests several ways in which sanitation-related bodily integrity, safety and security, privacy, and time may influence women’s mental health. For bodily integrity, which includes women’s choices and control over their bodies, studies have found that women restricted food and water and suppressed urination and defecation because of poor sanitation conditions, contributing to stress [10]. Similarly, women have reported a lack of sanitation-related privacy and safety, especially when openly defecating, as a source of anxiety, fear, and shame [10, 14, 15, 21–23]. Women have also reported feeling worried when they have had limited time for their sanitation needs and responsibilities [10, 14]. While sanitation-related bodily integrity, privacy, safety and security, and time are relevant to women’s mental health and well-being, there is limited quantitative research that demonstrates associations between these resources and mental health outcomes [17, 24].

This study aimed to investigate how sanitation-related resources – Bodily Integrity, Safety and Security, Privacy, and Time – are associated with key mental health outcomes (well-being, depression, and anxiety).

## 2 Methods

### 2.1 Study design

We conducted a secondary analysis of cross-sectional data collected for the Measuring Urban Sanitation and Empowerment (MUSE) project, which aimed to develop and validate measures of women’s sanitation-related empowerment in urban areas of low- and middle-income countries (LMICs) [25]. Surveys were conducted with women in eight cities across five countries in Asia and Africa: Meherpur and Saidpur, Bangladesh; Narsapur, Tiruchirappalli, and Warangal, India; Dakar, Senegal; Kampala, Uganda; and Lusaka, Zambia. Data collection occurred between 12/08/2021 and 20/05/2022 (Meherpur: 21/03/2022 - 09/04/2022; Saidpur: 21/03/2022 - 08/04/2022; Narsapur: 30/09/2021 - 25/11/2021; Tiruchirappalli: 10/03/2022 - 21/04/2022; Warangal: 12/08/2021 - 09/09/2021; Dakar: 11/04/2022 - 20/05/2022; Kampala: 11/04/2022 - 12/05/2022; Lusaka: 21/10/2021 - 18/11/2021). Further details about the study design were described in the MUSE protocol [25].

### 2.2 Participants and procedures

The study’s eight cities were purposively selected because of their partnership with the Citywide Inclusive Sanitation (CWIS) program. Neighborhoods in each city were also purposively selected in collaboration with CWIS partners and local officials, except in Bangladesh, where neighborhoods were randomly selected from a sampling frame of all neighborhoods in the two cities. A random walk technique was used to select households from within each neighborhood and an adult woman was recruited from each selected household. In-person surveys were implemented by trained female enumerators who were fluent in the local languages of each location. Surveys were conducted with 5,744 women (720 in Meherpur, 730 in Saidpur, 720 in Narsapur, 735 in Tiruchirappalli, 703 in Warangal, 709 in Dakar, 713 in Kampala, and 715 in Lusaka). Further information on participants and procedures is provided elsewhere [25, 26].

### 2.3 Measures

#### 2.3.1 Outcomes

We selected three outcomes – subjective mental well-being, depression, and anxiety – to assess different components of mental health. Scores for each outcome were calculated based on the measures’ guidelines, which note that scores should be used for screening and not diagnosis.

We used the WHO Well-being Index (WHO-5) to measure subjective mental well-being. The measure has five statements (e.g., my daily life has been filled with things that interest me) with response options that range from 0 (‘At no time’) to 5 (‘All of the time’). Summed scores range can range from 0-25, with lower scores indicating poorer well-being [27].

We used the two-item Patient Health Questionnaire (PHQ-2) to assess depression [28]. The PHQ-2 asks the frequency over the last two weeks of feeling ‘little interest or pleasure in doing things’ and ‘feeling down, depressed, or hopeless,’ with response options ranging from 0 (‘Not at all’) to 3 (‘Nearly every day’). The summed score can range from 0- 6 from normal to severe depression; a score of 3 or more indicates that major depressive disorder is likely [28].

We used the two-item Generalized Anxiety Disorder (GAD-2) measure to assess anxiety [29]. The GAD-2 includes two items that ask the past two-week frequency of ’feeling nervous, anxious, or on edge’ and ‘not being able to stop or control worrying.’ Response options range from 0 (‘Not at all’) to 3 (‘Nearly every day’). The summed score can range from 0-6 from normal to severe anxiety, and a score of 3 or more indicates that anxiety disorder is likely [29, 30].

#### 2.3.2 Exposures

The four primary exposures were sanitation-related empowerment resources – Bodily Integrity, Safety and Security, Privacy, and Time – which were measured using the validated Agency, Resources, and Institutional Structures for Sanitation-related Empowerment (ARISE) Scales [26]. ARISE definitions of Bodily Integrity, Safety and Security, Privacy, and Time can be found in **Table 1**. Resources scale scores were calculated as a simple, unweighted average of all items in each scale. The average scale scores can range from 0 to 4, with higher scores indicating greater sanitation-related empowerment resources [31].

**Table 1.** ARISE Scale definitions for Bodily Integrity, Safety and Security, Privacy, and Time.

| Scale<sup>1</sup> | Definition<sup>2</sup> | Factors & items<br>in scale | Response options | Score interpretation |
| --- | --- | --- | --- | --- |
| Bodily Integrity | Women's control over their bodies and ability to access and use their preferred sanitation location | 2 factors;<br>8 items | 1 (Never) to<br>4 (Always) | Higher scores indicate women have more frequent satisfaction with sanitation location and less frequent withholding and suppression |
| Safety and Security | Women's freedom from acts or threats of violence (physical or sexual), coercion, harassment, or force when accessing and using sanitation locations or engaging in sanitation-related decision-making processes in the public sphere | 5 factors;<br>18 items | 1 (Strongly Disagree) to<br>4 (Strongly Agree)<br>(For 3 factors)<br>1 (Never) to<br>4 (Always)<br>(For 2 factors) | Higher scores indicate women have lower perception of personal and other women's risk of sanitation-related harm |
| Privacy | Women's ability to maintain desired levels of privacy when accessing and | 1 factor;<br>5 items | 1 (Never) to<br>4 (Always) | Higher scores indicate that women have greater privacy for |

| utilizing sanitation locations |  |  |  | sanitation |
| --- | --- | --- | --- | --- |
| Time | Women's control over their time and labor spent on sanitation-related tasks and activities | 2 factors;<br>6 items | 1 (Strongly Disagree)<br>to<br>4 (Strongly Agree) | Higher scores indicate women have lower perceptions of sanitation needs and responsibilities as a burden on women's time |

<sup>1</sup> All scales include reverse-coded items
<sup>2</sup> Sanitation location is inclusive of sanitation facilities and open defecation

#### 2.3.3 Covariates

We made a priori decisions to include the following individual-level characteristics as covariates, based on theory and literature: life stage, self-rated physical health, highest level of completed schooling, and perceived social support [17, 32–37]. Four life stage categories were defined based on age and marital status: 1 (‘Unmarried, age 49 and younger’), 2 (‘Married under three years’), 3 (‘Married three years or more, age 49 and younger’), and 4 (‘Over 49 years of age, any marital status’)[32]. Self-rated physical health was measured by one question from the PROMIS global health subscale, which asks, “In general, how would you rate your physical health?” with response options ranging from 1 (‘Poor’) to 5 (‘Excellent’)[35]. Education completed (categorized as ‘primary or less’, ‘secondary’, and ‘post-secondary’) was used as a proxy for wealth [36, 37]. We assessed social support with the Multidimensional Scale for Perceived Social Support (MSPSS) [33]. This 12-item scale measures perceived social support from family, friends, and significant others [33]. We used the 8 items representing family and friends, as unmarried women were less likely to have a significant other [34]. Response options range from 0 (‘Completely Disagree’) to 4 (‘Completely Agree’), and the mean score can range from 0 to 4, with higher scores indicating greater perceived social support.

We also included sanitation environment characteristics as binary covariates: latrine sharing status (‘unshared’ or ‘shared’), lockable latrine, lighting along the way to latrine, lighting inside latrine, and physically challenging to access or use latrine [10, 14, 15].

### 2.4 Data Management and Analysis

We constructed an analytic dataset by pooling the data from Meherpur, Saidpur, Tiruchirappalli, Dakar, and Kampala. We excluded observations from Narsapur, Warangal, and Lusaka, because data were not collected in those three cities for Bodily Integrity. As noted above, we calculated scale scores for each of the four sub-domains of sanitation-related resources using all items in each scale; therefore, any observations with missing data for any item(s) within a scale were dropped. Additionally, the analytic sample for anxiety excluded observations from Dakar and Kampala because data were not collected in those two cities for the Multidimensional Scale of Perceived Social Support. Social support was included as a covariate in the anxiety model, because it was associated with anxiety in the pooled three-city model (Meherpur, Saidpur, Tiruchirappalli); however, social support was excluded as a covariate in the well-being and depression models, because it was not associated with either mental health outcome in the pooled three-city models.

Using the final analytic sample of all survey respondents with complete data (n = 2,122), we calculated univariate descriptive statistics for all study variables included in the analysis and then investigated associations between sanitation-related empowerment resources and mental health outcomes. We ran linear regressions of the WHO-5, PHQ-2, and GAD-2 scores on the four resources (Bodily Integrity, Safety and Security, Privacy, and Time) to calculate β coefficients. We used continuous outcome scale scores (vs. cut-off categorical scores) because we were interested in participant mental health beyond screening thresholds. For each regression analysis, we ran unadjusted and adjusted pooled models. Unadjusted models included the sanitation-related empowerment resources. Adjusted models included the sanitation-related empowerment resources, along with individual characteristics covariates and sanitation environment covariates. All models controlled for city and adjusted for neighborhood-level clustering. Individual city models are included in the **S2-S14 Tables.** We used Stata (version 18.0) for all analyses.

### 2.5 Ethics statement

Study protocols were reviewed and approved by ethics review committees in each site: Emory University in Atlanta, USA (IRB00110271), Makerere University in Kampala, Uganda (MAK-SHSREC Ref No. 2019-038), Comité National d’Ethique pour la Recherche en Santé in Dakar, Senegal (SEN21/85), International Institute of Health Management Research in New Delhi, India (IRB/2020-2021/001), and International Training Network-Bangladesh University of Engineering and Technology in Dhaka, Bangladesh (1.003/2022/02). Participants in all cities received compensation (either cash or in-kind) in accordance with local policies and ethical requirements. All women provided informed consent (written: Kampala, Dakar, Tiruchirappalli; verbal: Meherpur, Saidpur) before participation in the study, and verbal consent was witnessed and documented by enumerators.

## 3 Results

### 3.1 Sample size and socio-demographic characteristics

The analytic sample included 2,122 participants with a mean age of 35 years (SD = 11.5), and included 414 unmarried women (20%), 83 women married under three years (4%), 1,407 women married three years or more (66%), and 218 over age 49 with any marital status (10%). The largest proportion (49%) reported completing secondary school, 36% reported completing primary or less, and 15% reported completing post-secondary. The mean social support score was 3.1 (SD = 0.7), indicating high family and friend social support. A majority of the participants (71%) reported ‘good’, ‘very good’, or ‘excellent’ physical health. For sanitation environment, 69% reported having an unshared latrine, 92% reported being able to lock their latrine from the inside, 86% reported sufficient lighting inside their latrine, 93% reported sufficient lighting along the way to their latrine, and 10% reported that they experienced physical challenges with using their latrine (**Table 2).**

**Table 2.**
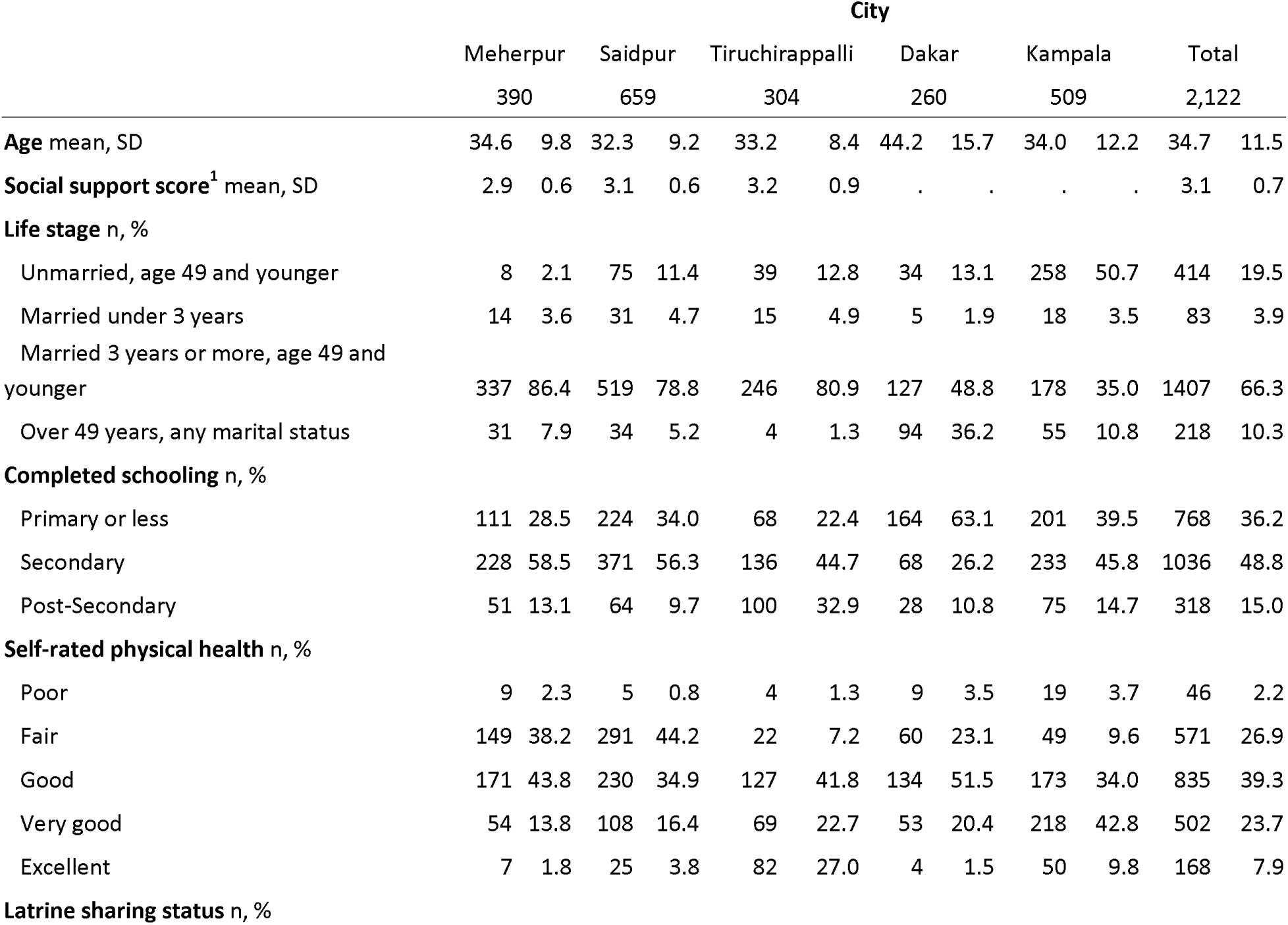
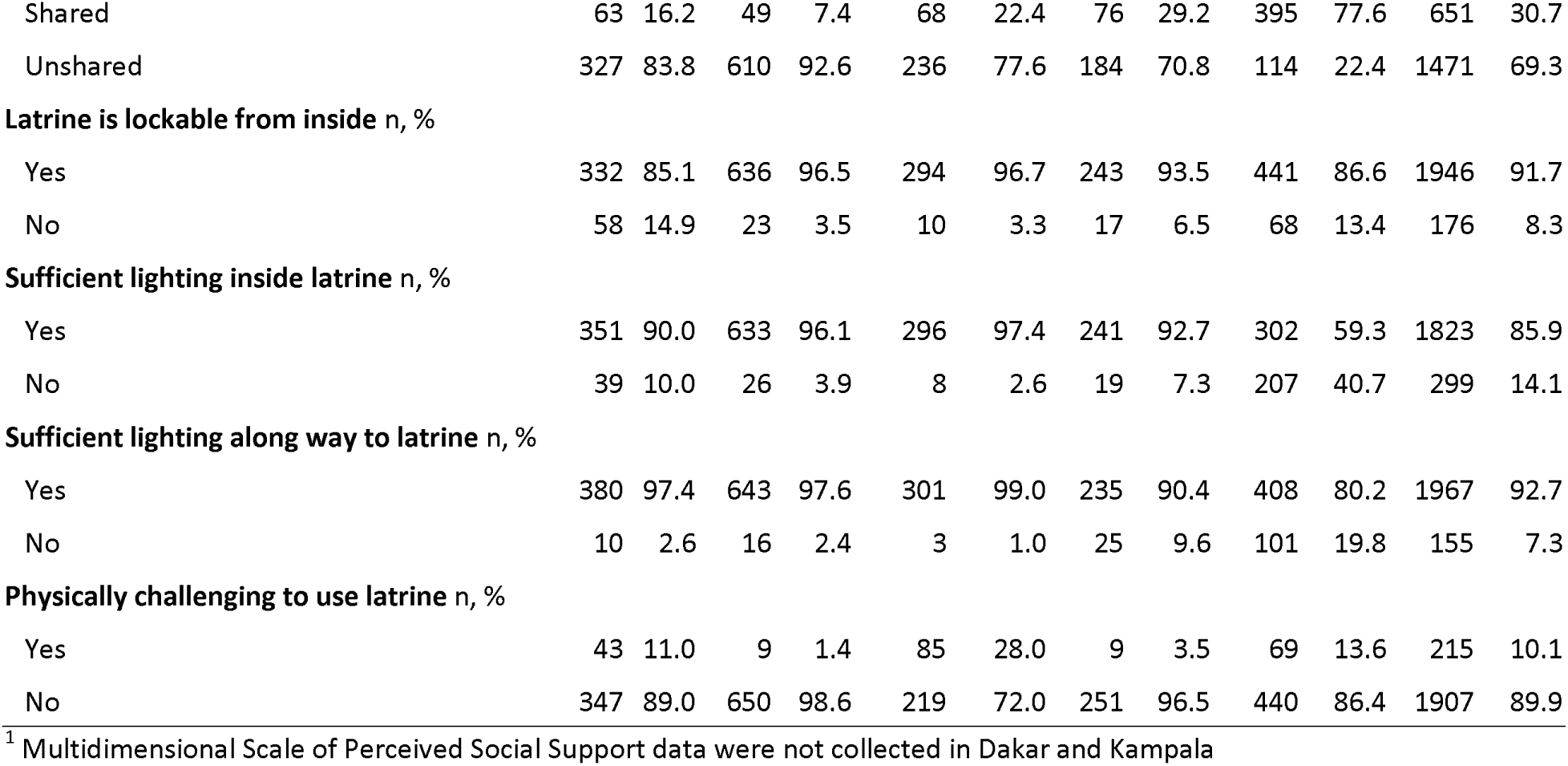
Demographic and sanitation environment characteristics of women respondents, by city (n = 2,122)

|  | City |  |  |  |  |  |  |  |  |  |  |  |
| --- | --- | --- | --- | --- | --- | --- | --- | --- | --- | --- | --- | --- |
|  | Meherpur |  | Saidpur |  | Tiruchirappalli |  | Dakar |  | Kampala |  | Total |  |
|  | 390 |  | 659 |  | 304 |  | 260 |  | 509 |  | 2,122 |  |
| **Age mean, SD** | 34.6 | 9.8 | 32.3 | 9.2 | 33.2 | 8.4 | 44.2 | 15.7 | 34.0 | 12.2 | 34.7 | 11.5 |
| **Social support score<sup>1</sup> mean, SD** | 2.9 | 0.6 | 3.1 | 0.6 | 3.2 | 0.9 | . | . | . | . | 3.1 | 0.7 |
| **Life stage n, %** |  |  |  |  |  |  |  |  |  |  |  |  |
| Unmarried, age 49 and younger | 8 | 2.1 | 75 | 11.4 | 39 | 12.8 | 34 | 13.1 | 258 | 50.7 | 414 | 19.5 |
| Married under 3 years | 14 | 3.6 | 31 | 4.7 | 15 | 4.9 | 5 | 1.9 | 18 | 3.5 | 83 | 3.9 |
| Married 3 years or more, age 49 and younger | 337 | 86.4 | 519 | 78.8 | 246 | 80.9 | 127 | 48.8 | 178 | 35.0 | 1407 | 66.3 |
| Over 49 years, any marital status | 31 | 7.9 | 34 | 5.2 | 4 | 1.3 | 94 | 36.2 | 55 | 10.8 | 218 | 10.3 |
| **Completed schooling n, %** |  |  |  |  |  |  |  |  |  |  |  |  |
| Primary or less | 111 | 28.5 | 224 | 34.0 | 68 | 22.4 | 164 | 63.1 | 201 | 39.5 | 768 | 36.2 |
| Secondary | 228 | 58.5 | 371 | 56.3 | 136 | 44.7 | 68 | 26.2 | 233 | 45.8 | 1036 | 48.8 |
| Post-Secondary | 51 | 13.1 | 64 | 9.7 | 100 | 32.9 | 28 | 10.8 | 75 | 14.7 | 318 | 15.0 |
| **Self-rated physical health n, %** |  |  |  |  |  |  |  |  |  |  |  |  |
| Poor | 9 | 2.3 | 5 | 0.8 | 4 | 1.3 | 9 | 3.5 | 19 | 3.7 | 46 | 2.2 |
| Fair | 149 | 38.2 | 291 | 44.2 | 22 | 7.2 | 60 | 23.1 | 49 | 9.6 | 571 | 26.9 |
| Good | 171 | 43.8 | 230 | 34.9 | 127 | 41.8 | 134 | 51.5 | 173 | 34.0 | 835 | 39.3 |
| Very good | 54 | 13.8 | 108 | 16.4 | 69 | 22.7 | 53 | 20.4 | 218 | 42.8 | 502 | 23.7 |
| Excellent | 7 | 1.8 | 25 | 3.8 | 82 | 27.0 | 4 | 1.5 | 50 | 9.8 | 168 | 7.9 |
| **Latrine sharing status n, %** |  |  |  |  |  |  |  |  |  |  |  |  |
| Shared | 63 | 16.2 | 49 | 7.4 | 68 | 22.4 | 76 | 29.2 | 395 | 77.6 | 651 | 30.7 |
| Unshared | 327 | 83.8 | 610 | 92.6 | 236 | 77.6 | 184 | 70.8 | 114 | 22.4 | 1471 | 69.3 |
| **Latrine is lockable from inside n, %** |  |  |  |  |  |  |  |  |  |  |  |  |
| Yes | 332 | 85.1 | 636 | 96.5 | 294 | 96.7 | 243 | 93.5 | 441 | 86.6 | 1946 | 91.7 |
| No | 58 | 14.9 | 23 | 3.5 | 10 | 3.3 | 17 | 6.5 | 68 | 13.4 | 176 | 8.3 |
| **Sufficient lighting inside latrine n, %** |  |  |  |  |  |  |  |  |  |  |  |  |
| Yes | 351 | 90.0 | 633 | 96.1 | 296 | 97.4 | 241 | 92.7 | 302 | 59.3 | 1823 | 85.9 |
| No | 39 | 10.0 | 26 | 3.9 | 8 | 2.6 | 19 | 7.3 | 207 | 40.7 | 299 | 14.1 |
| **Sufficient lighting along way to latrine n, %** |  |  |  |  |  |  |  |  |  |  |  |  |
| Yes | 380 | 97.4 | 643 | 97.6 | 301 | 99.0 | 235 | 90.4 | 408 | 80.2 | 1967 | 92.7 |
| No | 10 | 2.6 | 16 | 2.4 | 3 | 1.0 | 25 | 9.6 | 101 | 19.8 | 155 | 7.3 |
| **Physically challenging to use latrine n, %** |  |  |  |  |  |  |  |  |  |  |  |  |
| Yes | 43 | 11.0 | 9 | 1.4 | 85 | 28.0 | 9 | 3.5 | 69 | 13.6 | 215 | 10.1 |
| No | 347 | 89.0 | 650 | 98.6 | 219 | 72.0 | 251 | 96.5 | 440 | 86.4 | 1907 | 89.9 |

<sup>1</sup> Multidimensional Scale of Perceived Social Support data were not collected in Dakar and Kampala

### 3.2 Scores for well-being, depression, and anxiety

The overall mean scores for mental well-being were above the threshold of 13, indicating better well-being (mean = 17.2, SD = 5.8), though 19% (412) had well-being scores below 13, indicating poor well-being **(Fig 1 and Table 3)**. The overall mean scores for anxiety and depression were below the threshold of 3, indicating lower frequency of experiencing symptoms of both outcomes (depression mean = 1.1, SD = 1.4; anxiety mean = 1.0, SD = 1.4); however, 17% (356) had depression scores of 3 or more, indicating major depression disorder was likely and 12% (257) had anxiety scores of 3 or more, indicating anxiety disorder was likely **(Fig 1 and Table 3).** Well-being scores were moderately negatively correlated with anxiety and depression scores, and anxiety and depression scores were strongly positively correlated **(S1 Table).**

**Fig 1.**
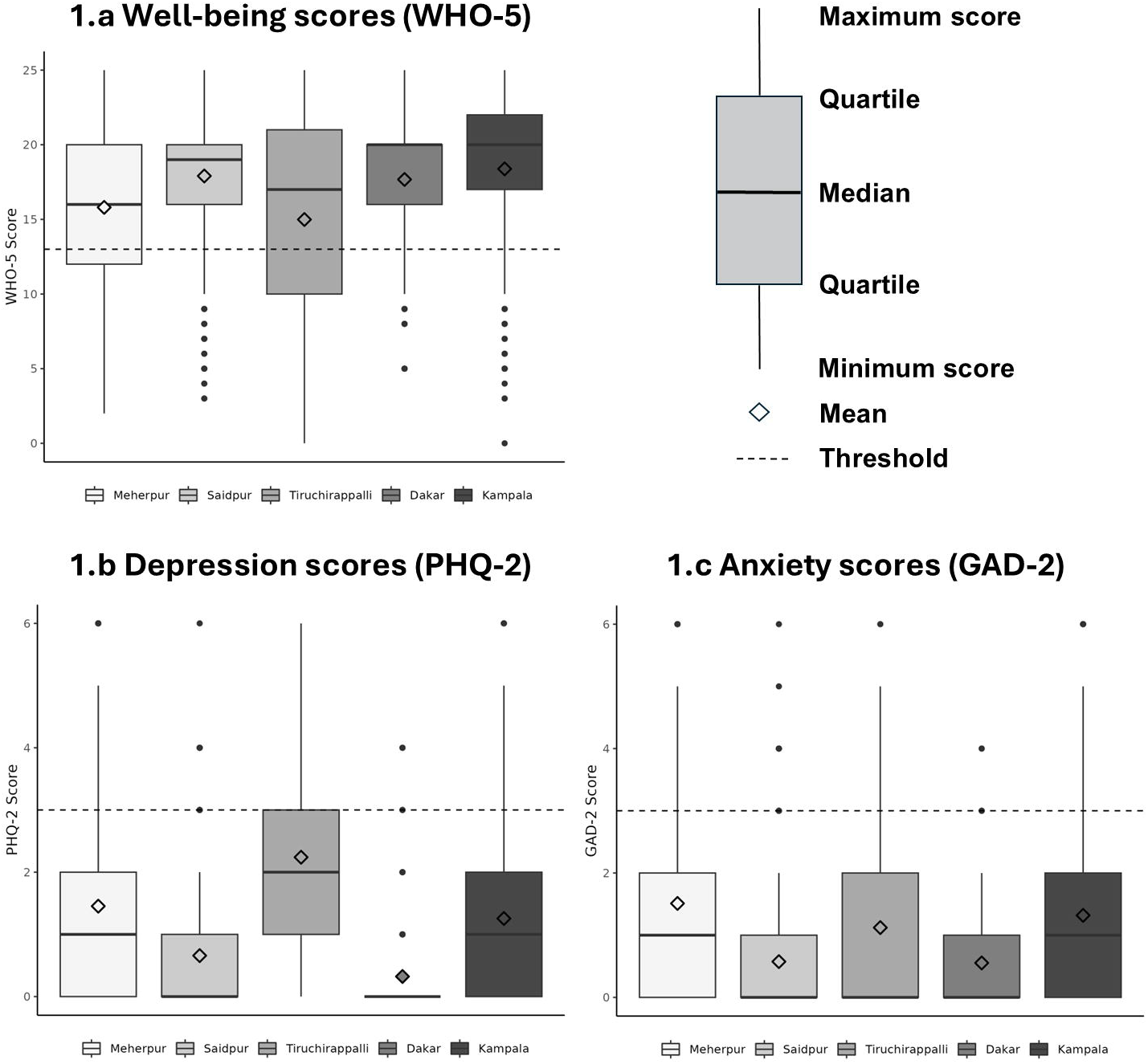
Boxplots of mental health outcome scores (well-being, depression, and anxiety), by city

**Table 3.**
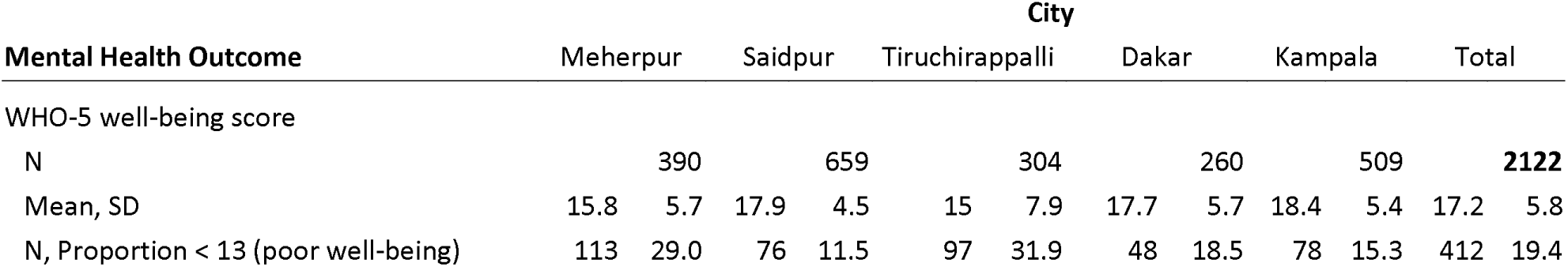
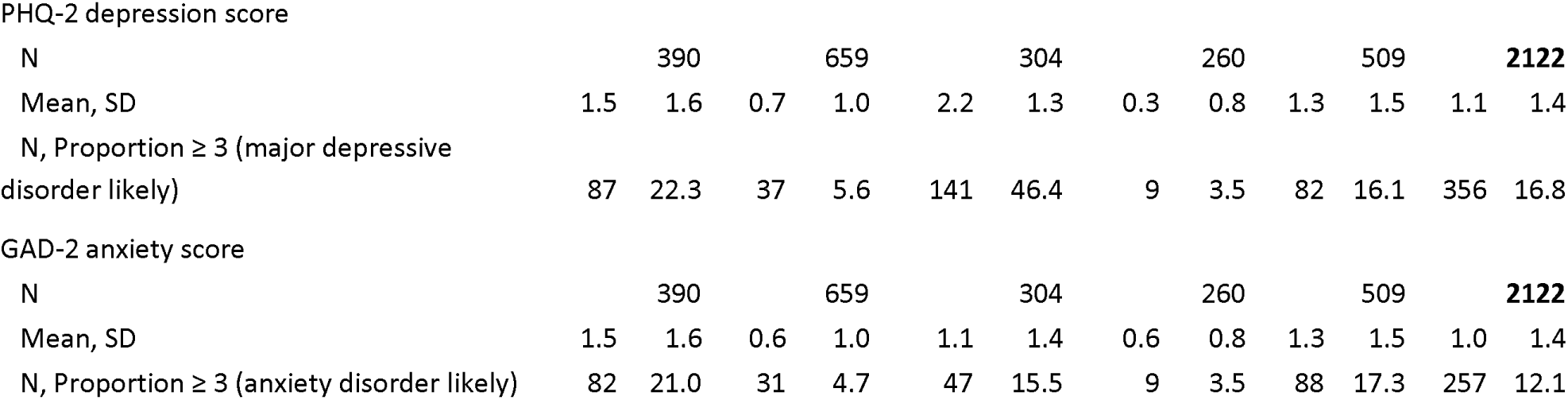
Mental health outcome scores (well-being, depression, and anxiety), by city.

### 3.3 Scores for sanitation-related Bodily Integrity, Safety and Security, Privacy, and Time

The overall scores for Bodily Integrity, Safety and Security, and Privacy were high and the overall score for Time was moderately high (Bodily Integrity mean = 3.7, SD = 0.4; Safety and Security mean = 3.5, SD = 0.4; Privacy mean = 3.8, SD = 0.5; Time mean = 3.2, SD = 0.5) (**Table 4)**.

**Table 4.** Sanitation-related empowerment resources scores of women respondents, by city.

| Resources | City |  |  |  |  |  |  |  |  |  |  |  |  |  |  |  |  |  |
| --- | --- | --- | --- | --- | --- | --- | --- | --- | --- | --- | --- | --- | --- | --- | --- | --- | --- | --- |
|  | Meherpur |  |  | Saidpur |  |  | Tiruchirappalli |  |  | Dakar |  |  | Kampala |  |  | Total |  |  |
|  | N | mean | SD | N | mean | SD | N | mean | SD | N | mean | SD | N | mean | SD | N | mean | SD |
| Bodily Integrity | 390 | 3.7 | 0.4 | 659 | 3.9 | 0.2 | 304 | 3.8 | 0.3 | 260 | 3.7 | 0.4 | 509 | 3.5 | 0.4 | **2122** | 3.7 | 0.4 |
| Safety and Security | 390 | 3.5 | 0.4 | 659 | 3.7 | 0.3 | 304 | 3.4 | 0.4 | 260 | 3.5 | 0.3 | 509 | 3.5 | 0.4 | **2122** | 3.5 | 0.4 |
| Privacy | 390 | 3.4 | 0.9 | 659 | 3.9 | 0.2 | 304 | 3.8 | 0.4 | 260 | 3.9 | 0.3 | 509 | 3.7 | 0.4 | **2122** | 3.8 | 0.5 |
| Time | 390 | 3.1 | 0.5 | 659 | 3.2 | 0.5 | 304 | 3.1 | 0.5 | 260 | 2.8 | 0.5 | 509 | 3.4 | 0.5 | **2122** | 3.2 | 0.5 |

### 3.4 Regression results

We found that higher scores for each of the four sub-domains of sanitation-related resources were associated with higher scores for well-being and lower scores for depression and anxiety. Individual city models were more mixed in their associations between the resources and mental health outcomes.

#### 3.4.1 Well-being

In the pooled adjusted well-being model, there was a positive association between Privacy and well-being (β = 2.0, p<.001), indicating that for each one-point increase in sanitation-related Privacy, well-being scores increased by two points. Well-being was not associated with Bodily Integrity (β = 0.9, p=.147), Safety and Security (β = 0.0, p=.964), or Time (β = 0.9, p=.084) **(Table 5).**

**Table 5.**
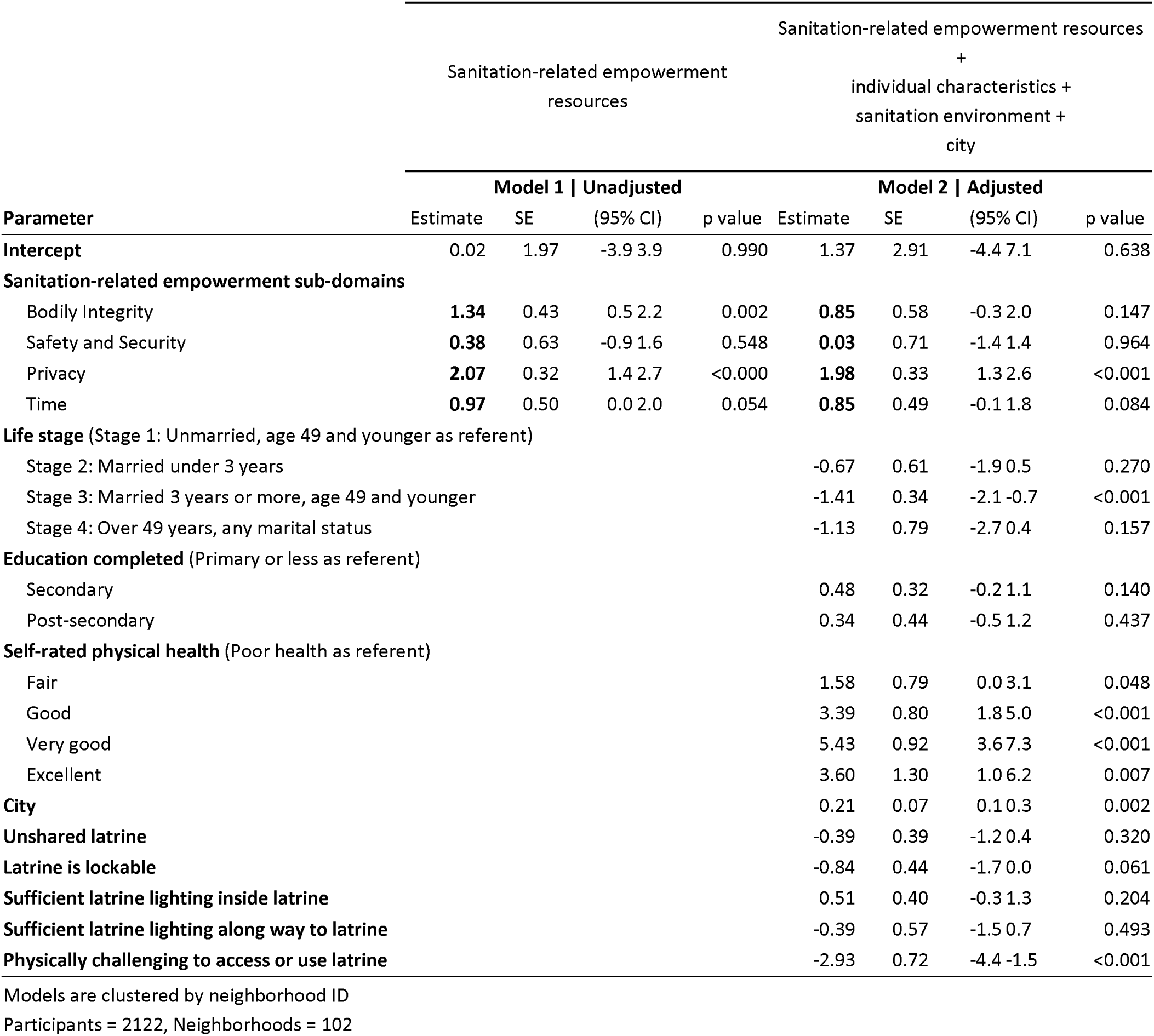
Association between sanitation-related empowerment resources sub-domains and well-being (WHO-5), in the pooled sample.

For individual-level characteristics, there was a negative association between women who were married for three years or more and their subjective well-being (β = -1.4, p<.001). Additionally, there were positive associations of self-rated physical health categories (fair through excellent, compared to poor health as referent) and well-being. For sanitation environment characteristics, there was a negative association between reporting physical challenges accessing or using latrines and well-being (β = -2.9, p<.001) **(Table 5)**.

In the individual city adjusted well-being models, we observed mixed associations between the four sub-domains and well-being. There were positive associations in two cities between Bodily Integrity and well-being (Saidpur β = 5.1, p<.001; Dakar β = 3.9, p=.011), as well as between Time and well-being (Saidpur β = 1.4, p=.022; Tiruchirappalli β = 5.9, p<.001), but no associations between either Bodily Integrity or Time and well-being in the other cities. In contrast, for Safety and Security, we observed a positive association with well-being in one city, Dakar (β = 5.0, p=.008), an inverse association in another city, Tiruchirappalli (β = -8.1, p<.001), and no association in the remaining three cities. Similarly, for Privacy, we observed positive associations in two cities (Meherpur β = 1.5, p<.001; Kampala β = 2.7, p<.001), an inverse association in one city, Dakar (β = -3.6, p<.001), and no associations in the remaining two cities **(S2-S6 Tables)**.

#### 3.4.2 Depression

In the pooled adjusted depression model, there were negative associations between Bodily Integrity and depression (β = -0.3, p=.002) and between Privacy and depression (β = -0.4, p<.001). Safety and Security was not associated with depression (β = 0.1, p=.578), nor was Time (β = -0.1, p=.110) **(Table 6)**.

**Table 6.**
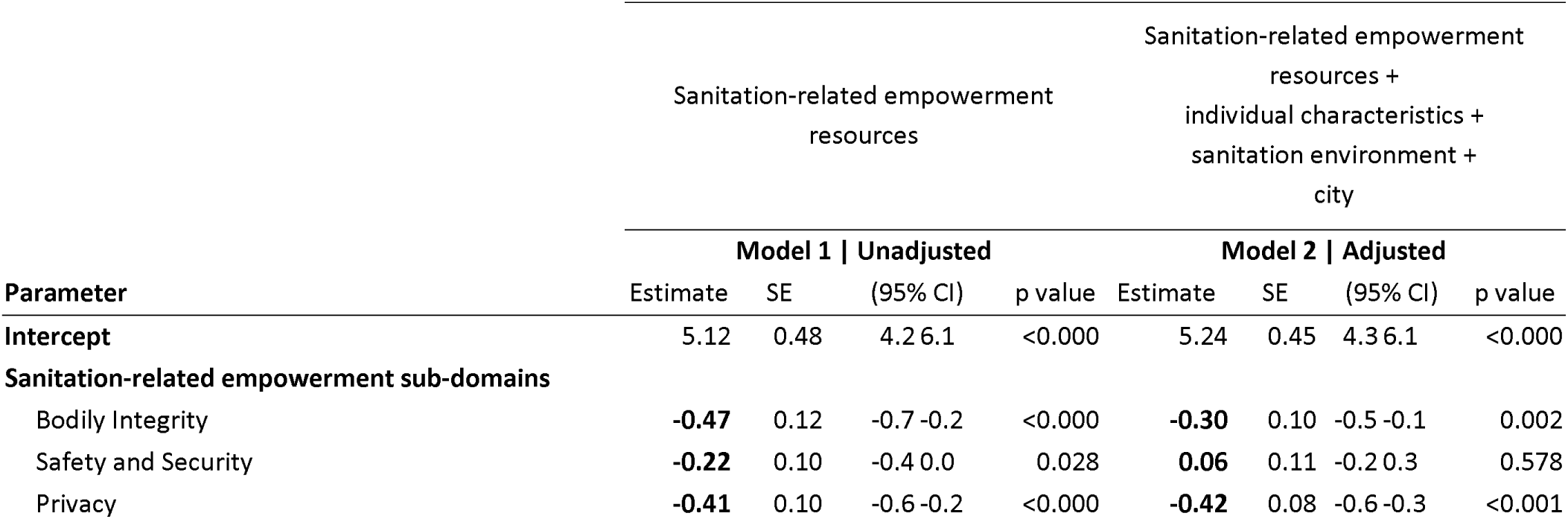
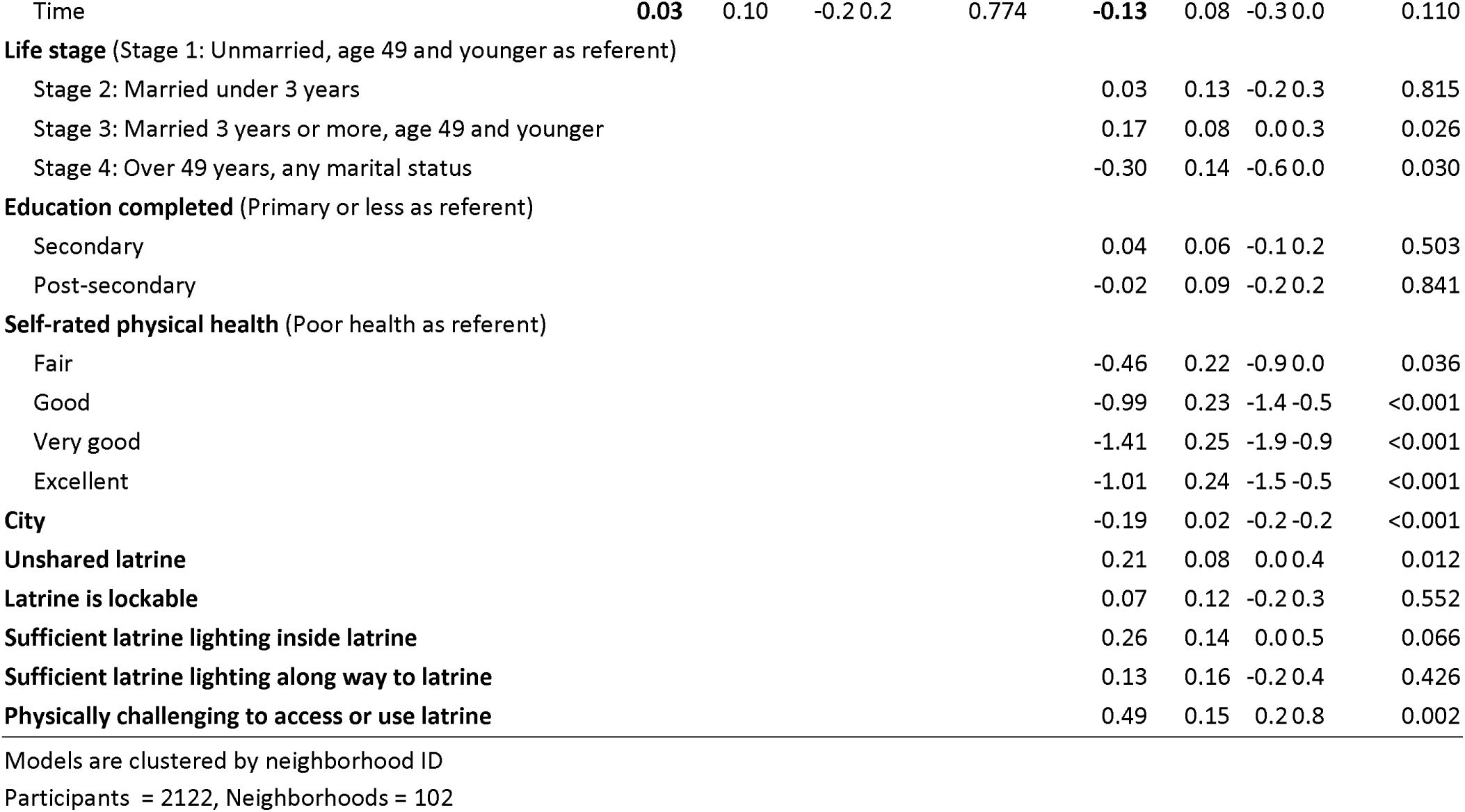
Association between sanitation-related empowerment resources sub-domains and depression (PHQ-2), in the pooled sample.

For individual-level characteristics, there were mixed associations for life stage, with a positive association between ‘stage 3: married three years or more’ and depression (β = 0.2, p=.026) and a negative association between ‘stage 4: over 49 years old’ and depression (β = -0.3, P=.030). All self-rated physical health categories (fair through excellent, with poor health as referent) were negatively associated with depression. For sanitation environment characteristics, there was a positive association between having an unshared latrine and depression (β = 0.2, p=.012). Additionally, experiencing physical challenges to access or use a latrine was positively associated with depression scores (β = 0.5, p=.002) **(Table 6)**.

In the individual city adjusted depression models, we observed fewer associations between the four sub-domains

and depression, compared to the results for well-being, but the observed associations were more consistent in their directionality. Specifically, there were negative associations between three of the sanitation-related empowerment resources (Bodily Integrity, Privacy, Time) and depression **(S7-S11 Tables)**. Bodily Integrity was negatively associated with depression in two of the five cities (Saidpur β = -1.1, p<.001; Dakar β = -0.3, p=.005). Similarly, Privacy was negatively associated with depression in two of the five cities (Dakar β = -0.9, p=.003; Kampala β = -0.8, p=.001). Time was negatively associated with depression in one city (Saidpur β = -0.4, p=.002). Safety and Security was not associated with depression in any of the five individual city models.

#### 3.4.3 Anxiety

In the pooled adjusted anxiety model, there were negative associations between all four sanitation-related empowerment resources and anxiety (Bodily Integrity β = -0.7, p<.001; Safety and Security β = -0.3, p=.025; Privacy β = -0.3, p=.037; Time β = -0.2, p=.009) **(Table 7)**.

**Table 7.**
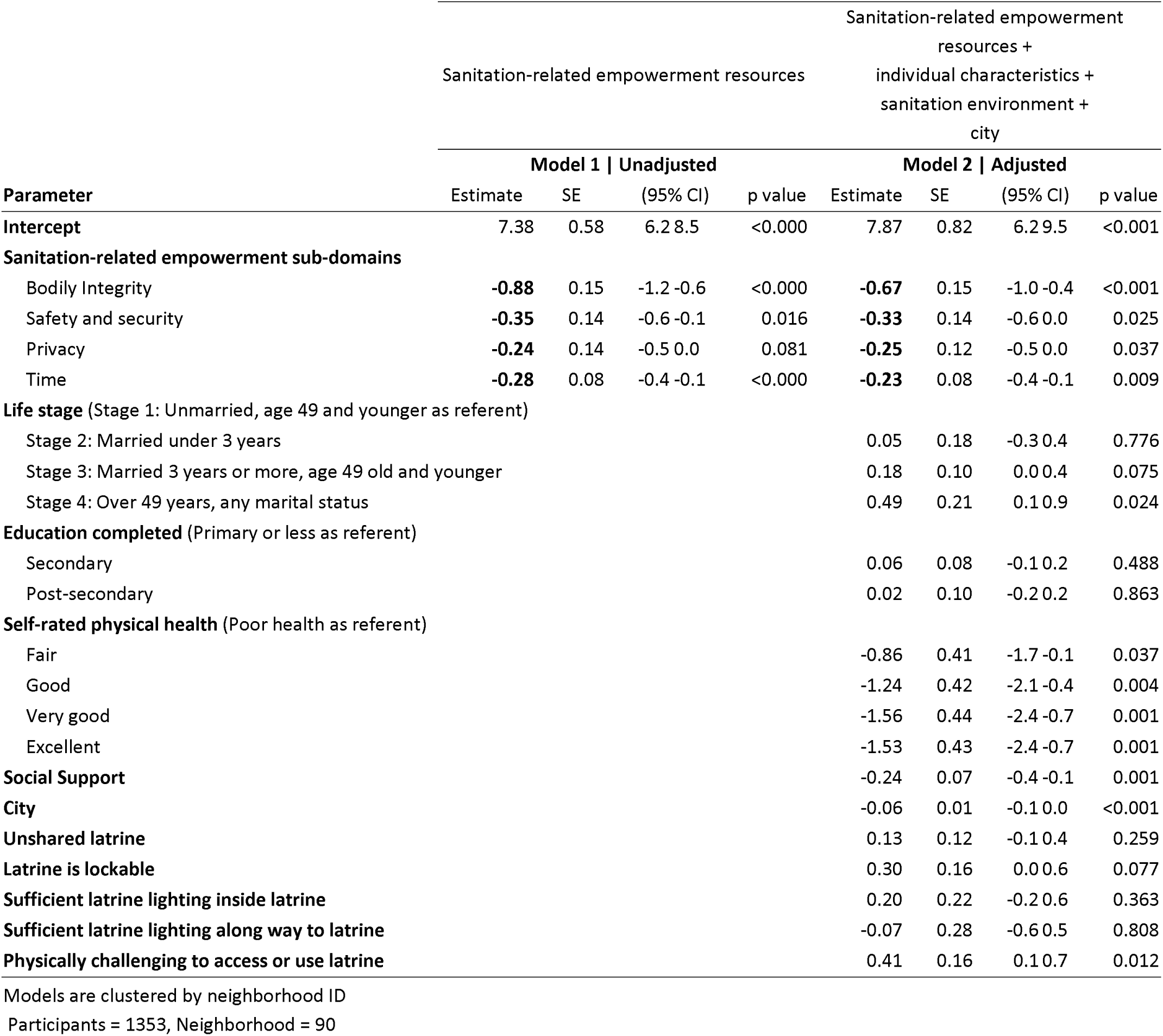
Association between sanitation-related empowerment resources sub-domains and anxiety (GAD-2), in the pooled sample.

For individual-level characteristics, there was a positive association between life stage (stage 4: over 49 years old, any marital status) and anxiety (β =0.5, p=.024). Additionally, self-rated physical health categories (fair through excellent, with poor health as referent) were negatively associated with anxiety, and social support scores were negatively associated with anxiety (β = -0.2, p=.001). For sanitation environment characteristics, experiencing physical challenges to access or use a latrine was positively associated with anxiety (β = 0.4, p=.012) **(Table 7)**.

In the individual city adjusted anxiety models, as with depression, there were negative associations between three

of the sanitation-related empowerment resources (Bodily integrity, Safety and Security, Time) and anxiety **(S12-S14 Tables)**. Bodily Integrity was negatively associated with anxiety in two of the three cities (Saidpur β = -0.9, p<.001; Tiruchirappalli β =-0.6, p=.030). Safety and Security was negatively associated with anxiety in one of the three cities (Tiruchirappalli β = -0.7, p=.001). Time was also negatively associated with anxiety in one city (Saidpur β = -0.5, p=.001). Unlike for well-being or depression, Privacy was not associated with anxiety in any of the three individual city models.

## 4 Discussion

This study examined relationships between sanitation-related empowerment resources (Bodily Integrity, Safety and Security, Privacy, and Time) and mental health outcomes (well-being, depression, and anxiety) among women in urban areas of Bangladesh, India, Senegal, and Uganda. Higher Privacy scores were associated with higher well-being scores (indicating better subjective mental well-being) and lower depression and anxiety scores (indicating lower frequency of experiencing symptoms of depression and anxiety). In addition, higher Bodily Integrity scores were associated with lower depression and anxiety scores. Notably, higher scores for all four sanitation-related resources (Bodily Integrity, Safety and Security, Privacy, and Time) were associated with lower anxiety scores. There was variation across city models, including mixed associations between resources and well-being.

Our study provides insights into women’s mental health in urban areas across multiple LMIC settings. Overall, while mean well-being scores were moderate and mean depression and anxiety scores were in the normal range, 19% of women had scores suggesting poor overall well-being, 17% had scores suggesting depression, and 12% had scores suggesting anxiety. Further, scores varied across cities, with greater proportions of women having scores suggesting poor mental health in Meherpur, Bangladesh and Tiruchirappalli, India. Globally, depression and anxiety are among the top ten leading causes of disease burden, and both disorders are 50% more common in women compared to men [38]. Leading mental health experts have called for further understanding of the causes of mental health, and although this study only looked at associations, our findings suggest that women’s sanitation-related resources may influence their mental health [38]. Our results provide support for these calls and reinforce the critical need for greater attention to women’s mental health, particularly in LMIC settings.

To our knowledge, this is the first study to examine relationships between sanitation-related Bodily Integrity and mental health outcomes. Across contexts, we found higher Bodily Integrity scores (women having greater control over their bodies and ability to access and use their preferred sanitation location) were associated with lower scores for both depression and anxiety, indicating lower frequency of symptoms of depression and anxiety. We also found sanitation- related Bodily Integrity to be associated with higher well-being in two cities (Saidpur and Dakar). Aspects of women’s bodily integrity, like suppressing urination and defecation urges and withholding food and water, have been increasingly discussed in sanitation research [18]. While these behaviors have been described as a form of bodily control, research demonstrates that women practice these behaviors to cope with unsupportive physical and social sanitation environments [7, 10, 14, 39–41]. As such, this exertion of bodily control is likely out of necessity, not preference, and may explain why Bodily Integrity was found to be associated with anxiety and depression across locations and with general well-being in two. For example, perceptions of sanitation facility cleanliness, privacy, safety, and adequacy (e.g., disposal bins for menstrual materials) can motivate suppression and withholding, as can social expectations, like deprioritizing personal needs to meet familial expectations (e.g., childcare, cooking), or limiting urination and defecation to specific times (e.g., dusk, dawn) to preserve dignity [10, 42–46]. A forthcoming study with women in Kampala, Uganda and Tiruchirappalli, India found that 93% of women in both populations reported suppression and 38% in Kampala and 16% in Tiruchirappalli reported withholding food and water; women’s negative perceptions of Privacy, Safety and Security, and their personal health were all associated with increased odds of withholding [47]. The link our study found between Bodily Integrity and mental health demonstrates that an inability to meet needs or be satisfied with sanitation environments are not merely unfortunate inconveniences, but issues that pose real risks to women that should be addressed.

Our finding that higher sanitation-related Privacy scores were associated with higher scores for well-being and lower scores for depression and anxiety adds to our understanding of the relationship between sanitation-related privacy and women’s mental health. Notably, we found Privacy scores to be associated with well-being and anxiety scores regardless of whether women had access to unshared sanitation facilities such as individual household latrines. This result aligns with a systematic review that found an absence of privacy influenced negative well-being and associated dimensions, like anxiety [13]. Sclar et al., identified that this pattern for privacy existed for women who practiced open defecation and for those who used unshared latrines, not only highlighting the importance of privacy across sanitation environments but emphasizing that access to an unshared facility can enable, but does not guarantee, privacy [13]. Our findings show that privacy is important to mental health, and that sanitation initiatives have an opportunity to improve women’s mental health by enhancing sanitation-related privacy.

The observed inverse association between sanitation-related Safety and Security scale scores and anxiety scores aligns with other research documenting women and girls’ fears related to safety and security threats when accessing sanitation. These included fear of harassment and non-partner physical and sexual violence, especially for those who practiced open defecation or used sanitation facilities outside the home [18]. These fears were often linked to features of sanitation facilities or locations such as lighting, lockability, and/or distance, and were especially intense for adolescent girls, young women, and minorities [8, 9, 14, 15, 18, 44, 48]. These types of fears may provoke anxiety more so than depression or overall poor well-being, which may explain why we did not observe an association between Safety and Security and the other two outcomes in our sample. In our study, individual city models indicated that the relationship between Safety and Security with anxiety was largely driven by the sample from Tiruchirappalli, which aligns with other research from India, specifically from urban slums in Bhubaneswar where women reported fear of sexual assault when accessing sanitation [14, 44]. Of the three cities in which we measured anxiety (Meherpur, Saidpur, and Tiruchirappalli), the percentage of respondents who used a shared latrine was highest in Tiruchirappalli, which may further contribute to the observed association.

This study also provides evidence that higher Time scores (indicating greater control over time and labor related to sanitation) were associated with lower anxiety scores. Studies have documented that women need to spend considerable time meeting personal sanitation needs, particularly if they need to travel to open defecation fields or shared latrines, as well as for household sanitation chores and caring for the sanitation needs of dependents [10, 41, 49–52]. Earlier qualitative research found that women have reported feeling anxiety and worry when they do not have access to latrines, have to queue at latrines, or have experienced conflict at home for taking too long to meet sanitation needs [14, 53, 54]. Women have also had to change their time of day for meeting sanitation needs to cope with a lack of privacy, safety, or access [10, 14]. The association between Time scores and anxiety scores suggests that time spent on sanitation needs and household responsibilities can be a burden and negatively affect women’s mental health.

### 4.1 Implications for research and practice

Our findings provide further evidence linking women’s sanitation-related experiences to mental health and is the latest study to find that access to sanitation infrastructure alone is not enough to meet the needs of women [12, 13, 17]. As such, sanitation initiatives need to move beyond a narrow focus on feces management and should strive to improve women’s experiences related to Bodily Integrity, Safety and Security, Privacy, and Time resources. Specifically, sanitation facilities should be intentionally designed, situated, operated, and maintained to enhance women’s experiences [15, 55, 56].

Existing guidance suggests design elements for female-friendly toilets, including proper lighting, lockable doors, and disposal bins [55, 56]. Other features like additional stalls in shared latrines to reduce queuing time and enough space for both women and dependents requiring toileting care, like children, may give women greater control over their time and labor spent on sanitation [55]. Similarly, guidance exists on gender-inclusive approaches that engage women to provide feedback and input on where and how facilities should be designed and placed [57]. Previous large-scale efforts that failed to include women in decision-making, placed household toilets in locations considered unacceptable, not private, and inconvenient, and so they were not used as a result [58]. Encouragingly, the recently published Priority Gender-Specific Indicators for WASH Monitoring Under SDG Targets 6.1 and 6.2 recommends a new set of indicators for national and global monitoring that assess perceptions of cleanliness, privacy, and safety at the individual level [59]. These indicators are meant to further motivate practitioners and policymakers to consider how they can influence women’s bodily integrity, safety and security, privacy, and time in sanitation programming.

Our study further demonstrates the value of measuring sub-domains of empowerment—in particular, Bodily Integrity, Safety and Security, Privacy, and Time—as independent constructs, to understand gender-specific experiences related to sanitation in urban settings. For example, as noted above, in our pooled regression models, Privacy was the only one of the four sub-domains included in our study that was significantly associated with all three mental health outcomes (well-being, depression, and anxiety). While scale scores for Privacy were generally high across our study populations in the five cities, there were still women with privacy concerns. Future research and programs could use the ARISE Privacy scale, for example, to identify populations (or sub-populations) with lower Privacy scores and target these populations for future privacy-related infrastructural improvements. We recommend using the ARISE scales before sanitation programming begins to assess baseline needs and to measure changes in women’s mental wellbeing during and after programs end. The ARISE scales can also be used to examine causal relationships between programs, sub-domains of empowerment, and health and well-being outcomes for women. While the ARISE scales are designed for urban settings, other validated tools exist for rural settings, such as the WASH-GEM, which can be used to measure similar constructs [60].

### 4.2 Strengths and limitations

This work contributes to a nascent body of research identifying linkages between sanitation-related experiences and mental health outcomes. Our study leverages data from urban contexts in four countries, demonstrating that relationships between sanitation-related resources and mental well-being are not limited to specific contexts or settings. In addition, the research uses measures of sanitation-related empowerment that have been developed and rigorously validated for use across urban contexts in Africa and South Asia, allowing for the consistent measurement of sub-domains of empowerment across populations.

This study also has several limitations, which should be considered when interpreting the results. Our sample did not include non-binary or transgender populations, as well as women sanitation workers, who have different sanitation and mental health experiences [61–63]. Our analysis did not include several variables that may be associated with mental health, such as wealth or housing quality [64]. While we were not able to control for wealth and assets in our models, we included a variable for highest completed education in our analysis, as more years of education have been shown to have a protective effect on mortality across economic contexts [37]. Additionally, we note that mental health measures have been predominantly developed based on Western medicine-derived constructs [65, 66]. Our assessments of well-being, depression, and anxiety were not based on local perceptions of these outcomes; rather, we used the previously validated WHO-5, PHQ-2, and GAD-2 measures.

## 5 Conclusion

Overall, our findings support existing calls for a more comprehensive view of sanitation, which considers the intersectional needs of women. Our results demonstrate important linkages between women’s sanitation-related resources of Bodily Integrity, Safety and Security, Privacy, and Time and mental health. Thus, sanitation infrastructure should include features to improve women’s bodily integrity, safety and security, privacy, and control over time. Evaluations of sanitation interventions, programs, and environments should incorporate assessments of sanitation-related resources and mental health outcomes for comprehensive assessments of impact beyond mere infrastructure availability. Sanitation-users’ perceptions of their resources are crucial, particularly for women. It is important to center their experiences in intervention design and policymaking to ensure that sanitation investments contribute to – and do not detract from – overall well-being.

## Data Availability

[DOI will be made available after acceptance.]

## Acknowledgements

We are grateful to Deepa Karthykeyan, Kun Zhang, Arjun Sharma, and Josephine Goma of Athena Infonomics; Shravya Narakula, Thejasweeni Bingisetty, and Vimalan Karunakaran of Civic Fulcrum; Catherine Mwanje of CME Solutions; Amadou Ba of People and Data; Alauddin Ahmed, Makfe Farah, Rakib Uddin Ahmed, and Samina of ITN-BUET; Kazi Amin of WaterAid Bangladesh; Tahmid Hossain of Practical Action; and Shaila Shahid of the Department of Public Health Engineering of the Bangladesh government for support, coordination, and supervision of study activities. We extend our gratitude to all members of the data collection teams in each city. We are also grateful to the Citywide Inclusive Sanitation (CWIS) partners for their input and support, especially Allan Nkurunziza and Hilda Sande Kwesiga of Kampala Capital City Authority. We are grateful to all study participants for sharing their time and contributing to the research.

## Supporting information captions

**S1 - S14 Tables**

