## Supplemental Tables for "Sanitation-related empowerment resources are associated with women’s well-being, anxiety, and depression: findings from Bangladesh, India, Senegal, and Uganda"

**Supporting Information Table of Contents**

| **Table S1.** Polychoric correlations of mental health outcome scores | 2 |
| --- | --- |
| **Table S2.** Association between sanitation-related empowerment resources sub-domains and **well-being (WHO-5)**, in the Meherpur sample (n = 390) | 3 |
| **Table S3.** Association between sanitation-related empowerment resources sub-domains and **well-being (WHO-5)**, in the Saidpur sample (n = 659) | 5 |
| **Table S4.** Association between sanitation-related empowerment resources sub-domains and **well-being (WHO-5)**, in the Tiruchirappalli sample (n = 304) | 7 |
| **Table S5.** Association between sanitation-related empowerment resources sub-domains and **well-being (WHO-5)**, in the Dakar sample (n = 260) | 9 |
| **Table S6.** Association between sanitation-related empowerment resources sub-domains and **well-being (WHO-5)**, in the Kampala sample (n = 509) | 11 |
| **Table S7.** Association between sanitation-related empowerment resources sub-domains and **depression (PHQ-2)**, in the Meherpur sample (n = 390) | 13 |
| **Table S8.** Association between sanitation-related empowerment resources sub-domains and **depression (PHQ-2)**, in the Saidpur sample (n = 659) | 15 |
| **Table S9.** Association between sanitation-related empowerment resources sub-domains and **depression (PHQ-2)**, in the Tiruchirappalli sample (n = 304) | 17 |
| **Table S10.** Association between sanitation-related empowerment resources sub-domains and **depression (PHQ-2)**, in the Dakar sample (n = 260) | 19 |
| **Table S11.** Association between sanitation-related empowerment resources sub-domains and **depression (PHQ-2)**, in the Kampala sample (n = 509) | 21 |
| **Table S12.** Association between sanitation-related empowerment resources sub-domains and **anxiety (GAD-2)**, in the Meherpur sample (n = 390) | 23 |
| **Table S13.** Association between sanitation-related empowerment resources sub-domains and **anxiety (GAD-2)**, in the Saidpur sample (n = 659) | 25 |
| **Table S14.** Association between sanitation-related empowerment resources sub-domains and **anxiety (GAD-2)**, in the Tiruchirappalli sample (n = 304) | 27 |

**Sanitation-related empowerment resources are associated with women’s well-being, anxiety, and depression: findings from Bangladesh, India, Senegal, and Uganda**

Thea Mink, Madeleine Patrick, Amelia Conrad, Tanvir Ahmed, Srishty Arun, Vinod Ramanarayanan, Niladri Chakraborti, Y. Malini Reddy, Abhilaasha Nagarajan, Tanushree Bhan, Sheela S. Sinharoy, Bethany A. Caruso

**Table S1.** Polychoric correlations of mental health outcome scores

|  | **1. Well-being** | **2. Depression** |
| --- | --- | --- |
| **1. Well-being** |  |  |
| **2. Depression** | -0.471 |  |
| **3. Anxiety** | -0.425 | 0.745 |

**Sanitation-related empowerment resources are associated with women’s well-being, anxiety, and depression: findings from Bangladesh, India, Senegal, and Uganda**

Thea Mink, Madeleine Patrick, Amelia Conrad, Tanvir Ahmed, Srishty Arun, Vinod Ramanarayanan, Niladri Chakraborti, Y. Malini Reddy, Abhilaasha Nagarajan, Tanushree Bhan, Sheela S. Sinharoy, Bethany A. Caruso

| **Table S2.** Association between sanitation-related empowerment resources sub-domains and **well-being (WHO-5)**, in the Meherpur sample (n = 390) | | | | | | | | | | |
| --- | --- | --- | --- | --- | --- | --- | --- | --- | --- | --- |
|  | Sanitation-related empowerment resources | | | | | Sanitation-related empowerment resources +  individual characteristics +  sanitation environment +  city | | | | |
|  | **Model 1 \| Unadjusted** | | | | | **Model 3 \| Adjusted** | | | | |
| **Parameter** | Estimate | SE | (95% CI) | | p value | Estimate | SE | (95% CI) | | p value |
| **Intercept** | -0.94 | 3.81 | -8.7 | 6.8 | 0.807 | 4.44 | 5.90 | -7.6 | 16.4 | 0.458 |
| **Sanitation-related empowerment sub-domains** |  |  |  |  |  |  |  |  |  |  |
| Bodily Integrity | **1.62** | 1.02 | -0.5 | 3.7 | 0.121 | **0.39** | 1.08 | -1.8 | 2.6 | 0.722 |
| Safety and Security | **2.60** | 1.06 | 0.4 | 4.8 | 0.020 | **1.90** | 1.00 | -0.1 | 3.9 | 0.067 |
| Privacy | **1.14** | 0.42 | 0.3 | 2.0 | 0.011 | **1.52** | 0.35 | 0.8 | 2.2 | <0.001 |
| Time | **-0.83** | 0.64 | -2.1 | 0.5 | 0.207 | **-1.02** | 0.68 | -2.4 | 0.4 | 0.143 |
| **Life stage** (Stage 1: Unmarried, age 49 and younger as referent) | |  |  |  |  |  |  |  |  |  |
| Stage 2: Married under 3 years |  |  |  |  |  | 0.56 | 1.70 | -2.9 | 4.0 | 0.744 |
| Stage 3: Married 3 years or more, age 49 and younger |  |  |  |  |  | -1.21 | 1.42 | -4.1 | 1.7 | 0.397 |
| Stage 4: Over 49 years old, any marital status |  |  |  |  |  | -1.52 | 1.62 | -4.8 | 1.8 | 0.355 |
| **Education completed** (primary or less as referent) |  |  |  |  |  |  |  |  |  |  |
| Secondary |  |  |  |  |  | 1.04 | 0.69 | -0.4 | 2.4 | 0.140 |
| Post-secondary |  |  |  |  |  | 0.41 | 0.86 | -1.3 | 2.2 | 0.635 |
| **Self-rated physical health** (poor health as referent) |  |  |  |  |  |  |  |  |  |  |
| Fair |  |  |  |  |  | -0.63 | 1.56 | -3.8 | 2.5 | 0.690 |
| Good |  |  |  |  |  | 0.37 | 1.54 | -2.8 | 3.5 | 0.814 |
| Very good |  |  |  |  |  | 3.22 | 1.77 | -0.4 | 6.8 | 0.078 |
| Excellent |  |  |  |  |  | 5.81 | 1.98 | 1.8 | 9.8 | 0.006 |
| **Unshared latrine** |  |  |  |  |  | -0.26 | 0.76 | -1.8 | 1.3 | 0.731 |
| **Latrine is lockable** |  |  |  |  |  | -1.02 | 0.79 | -2.6 | 0.6 | 0.209 |
| **Sufficient latrine lighting inside latrine** |  |  |  |  |  | 1.61 | 0.94 | -0.3 | 3.5 | 0.096 |
| **Sufficient latrine lighting along way to latrine** |  |  |  |  |  | 1.02 | 1.22 | -1.5 | 3.5 | 0.411 |
| **Physically challenging to access or use latrine** |  |  |  |  |  | -1.17 | 0.85 | -2.9 | 0.6 | 0.179 |
| Models are clustered by neighborhood (n = 34) | | | | | | | | | | |

**Sanitation-related empowerment resources are associated with women’s well-being, anxiety, and depression: findings from Bangladesh, India, Senegal, and Uganda**

Thea Mink, Madeleine Patrick, Amelia Conrad, Tanvir Ahmed, Srishty Arun, Vinod Ramanarayanan, Niladri Chakraborti, Y. Malini Reddy, Abhilaasha Nagarajan, Tanushree Bhan, Sheela S. Sinharoy, Bethany A. Caruso

| **Table S3.** Association between sanitation-related empowerment resources sub-domains and **well-being (WHO-5)**, in the Saidpur sample (n = 659) | | | | | | | | | | |
| --- | --- | --- | --- | --- | --- | --- | --- | --- | --- | --- |
|  | Sanitation-related empowerment resources | | | | | Sanitation-related empowerment resources +  individual characteristics +  sanitation environment +  city | | | | |
|  | **Model 1 \| Unadjusted** | | | | | **Model 3 \| Adjusted** | | | | |
| **Parameter** | Estimate | SE | (95% CI) | | p value | Estimate | SE | (95% CI) | | p value |
| **Intercept** | -8.12 | 3.87 | -15.9 | -0.3 | 0.042 | -15.54 | 6.89 | -29.5 | -1.6 | 0.029 |
| **Sanitation-related empowerment sub-domains** |  |  |  |  |  |  |  |  |  |  |
| Bodily Integrity | **5.30** | 0.95 | 3.4 | 7.2 | <0.000 | **5.15** | 1.25 | 2.6 | 7.7 | <0.001 |
| Safety and Security | **0.05** | 1.10 | -2.2 | 2.3 | 0.966 | **0.48** | 1.08 | -1.7 | 2.7 | 0.657 |
| Privacy | **-0.49** | 0.91 | -2.3 | 1.3 | 0.589 | **1.07** | 1.11 | -1.2 | 3.3 | 0.340 |
| Time | **2.30** | 0.56 | 1.2 | 3.4 | <0.000 | **1.35** | 0.57 | 0.2 | 2.5 | 0.022 |
| **Life stage** (Stage 1: Unmarried, age 49 and younger as referent) | |  |  |  |  |  |  |  |  |  |
| Stage 2: Married under 3 years |  |  |  |  |  | -0.07 | 0.88 | -1.9 | 1.7 | 0.933 |
| Stage 3: Married 3 years or more, age 49 and younger |  |  |  |  |  | 0.22 | 0.46 | -0.7 | 1.2 | 0.637 |
| Stage 4: Over 49 years, any marital status |  |  |  |  |  | -0.56 | 0.74 | -2.1 | 0.9 | 0.449 |
| **Education completed** (primary or less as referent) |  |  |  |  |  |  |  |  |  |  |
| Secondary |  |  |  |  |  | 0.96 | 0.40 | 0.1 | 1.8 | 0.022 |
| Post-secondary |  |  |  |  |  | 1.30 | 0.60 | 0.1 | 2.5 | 0.034 |
| **Self-rated physical health** (poor health as referent) |  |  |  |  |  |  |  |  |  |  |
| Fair |  |  |  |  |  | 3.34 | 1.55 | 0.2 | 6.5 | 0.036 |
| Good |  |  |  |  |  | 5.82 | 1.37 | 3.0 | 8.6 | <0.001 |
| Very good |  |  |  |  |  | 6.62 | 1.40 | 3.8 | 9.5 | <0.001 |
| Excellent |  |  |  |  |  | 5.22 | 1.64 | 1.9 | 8.5 | 0.003 |
| **Unshared latrine** |  |  |  |  |  | 0.27 | 0.68 | -1.1 | 1.6 | 0.689 |
| **Latrine is lockable** |  |  |  |  |  | -3.04 | 0.70 | -4.4 | -1.6 | <0.001 |
| **Sufficient latrine lighting inside latrine** |  |  |  |  |  | 0.37 | 0.94 | -1.5 | 2.3 | 0.695 |
| **Sufficient latrine lighting along way to latrine** |  |  |  |  |  | -0.11 | 0.85 | -1.8 | 1.6 | 0.900 |
| **Physically challenging to access or use latrine** |  |  |  |  |  | 0.51 | 1.40 | -2.3 | 3.3 | 0.717 |
| Models are clustered by neighborhood (n = 43) | | | | | | | | | | |

**Sanitation-related empowerment resources are associated with women’s well-being, anxiety, and depression: findings from Bangladesh, India, Senegal, and Uganda**

Thea Mink, Madeleine Patrick, Amelia Conrad, Tanvir Ahmed, Srishty Arun, Vinod Ramanarayanan, Niladri Chakraborti, Y. Malini Reddy, Abhilaasha Nagarajan, Tanushree Bhan, Sheela S. Sinharoy, Bethany A. Caruso

| **Table S4.** Association between sanitation-related empowerment resources sub-domains and **well-being (WHO-5)**, in the Tiruchirappalli sample (n = 304) | | | | | | | | | | |
| --- | --- | --- | --- | --- | --- | --- | --- | --- | --- | --- |
|  | Sanitation-related empowerment resources | | | | | Sanitation-related empowerment resources +  individual characteristics +  sanitation environment +  city | | | | |
|  | **Model 1 \| Unadjusted** | | | | | **Model 3 \| Adjusted** | | | | |
| **Parameter** | Estimate | SE | (95% CI) | | p value | Estimate | SE | (95% CI) | | p value |
| **Intercept** | 7.88 | 2.53 | 2.4 | 13.4 | 0.009 | 12.85 | 6.81 | -2.0 | 27.7 | 0.083 |
| **Sanitation-related empowerment sub-domains** |  |  |  |  |  |  |  |  |  |  |
| Bodily Integrity | 1.19 | 0.85 | -0.7 | 3.0 | 0.185 | **0.62** | 0.96 | -1.5 | 2.7 | 0.528 |
| Safety and Security | -8.85 | 1.36 | -11.8 | -5.9 | <0.000 | **-8.14** | 1.26 | -10.9 | -5.4 | <0.001 |
| Privacy | 2.54 | 0.89 | 0.6 | 4.5 | 0.015 | **2.73** | 1.36 | -0.2 | 5.7 | 0.067 |
| Time | 7.53 | 0.98 | 5.4 | 9.7 | <0.000 | **5.86** | 1.13 | 3.4 | 8.3 | <0.001 |
| **Life stage** (Stage 1: Unmarried, age 49 and younger as referent) | |  |  |  |  |  |  |  |  |  |
| Stage 2: Married under 3 years |  |  |  |  |  | -1.98 | 1.53 | -5.3 | 1.3 | 0.219 |
| Stage 3: Married 3 years or more, age 49 and younger |  |  |  |  |  | -1.59 | 0.99 | -3.8 | 0.6 | 0.134 |
| Stage 4: Over 49 years, any marital status |  |  |  |  |  | -2.12 | 2.45 | -7.5 | 3.2 | 0.403 |
| **Education completed** (primary or less as referent) |  |  |  |  |  |  |  |  |  |  |
| Secondary |  |  |  |  |  | -1.94 | 1.10 | -4.3 | 0.5 | 0.103 |
| Post-secondary |  |  |  |  |  | -2.33 | 0.81 | -4.1 | -0.6 | 0.014 |
| **Self-rated physical health** (poor health as referent) |  |  |  |  |  |  |  |  |  |  |
| Fair |  |  |  |  |  | -1.27 | 2.44 | -6.6 | 4.0 | 0.611 |
| Good |  |  |  |  |  | -1.35 | 2.17 | -6.1 | 3.4 | 0.545 |
| Very good |  |  |  |  |  | 1.77 | 2.26 | -3.1 | 6.7 | 0.448 |
| Excellent |  |  |  |  |  | -1.67 | 2.32 | -6.7 | 3.4 | 0.484 |
| **Unshared latrine** |  |  |  |  |  | 0.69 | 1.00 | -1.5 | 2.9 | 0.502 |
| **Latrine is lockable** |  |  |  |  |  | 5.85 | 1.41 | 2.8 | 8.9 | 0.001 |
| **Sufficient latrine lighting inside latrine** |  |  |  |  |  | 0.40 | 5.72 | -12.1 | 12.9 | 0.945 |
| **Sufficient latrine lighting along way to latrine** |  |  |  |  |  | -2.41 | 5.72 | -14.9 | 10.1 | 0.680 |
| **Physically challenging to access or use latrine** |  |  |  |  |  | -4.80 | 0.58 | -6.1 | -3.5 | <0.001 |
| Models are clustered by neighborhood (n = 13) |  |  |  |  |  |  |  |  |  |  |

**Sanitation-related empowerment resources are associated with women’s well-being, anxiety, and depression: findings from Bangladesh, India, Senegal, and Uganda**

Thea Mink, Madeleine Patrick, Amelia Conrad, Tanvir Ahmed, Srishty Arun, Vinod Ramanarayanan, Niladri Chakraborti, Y. Malini Reddy, Abhilaasha Nagarajan, Tanushree Bhan, Sheela S. Sinharoy, Bethany A. Caruso

| **Table S5.** Association between sanitation-related empowerment resources sub-domains and **well-being (WHO-5)**, in the Dakar sample (n = 260) | | | | | | | | | | |
| --- | --- | --- | --- | --- | --- | --- | --- | --- | --- | --- |
|  | Sanitation-related empowerment resources | | | | | Sanitation-related empowerment resources +  individual characteristics +  sanitation environment +  city | | | | |
|  | **Model 1 \| Unadjusted** | | | | | **Model 3 \| Adjusted** | | | | |
| **Parameter** | Estimate | SE | (95% CI) | | p value | Estimate | SE | (95% CI) | | p value |
| **Intercept** | -6.72 | 3.37 | -14.5 | 1.0 | 0.081 | 0.76 | 3.97 | -8.6 | 10.2 | 0.853 |
| **Sanitation-related empowerment sub-domains** |  |  |  |  |  |  |  |  |  |  |
| Bodily Integrity | **2.49** | 1.82 | -1.7 | 6.7 | 0.209 | **3.94** | 1.14 | 1.2 | 6.6 | 0.011 |
| Safety and Security | **4.61** | 2.11 | -0.3 | 9.5 | 0.060 | **5.02** | 1.38 | 1.7 | 8.3 | 0.008 |
| Privacy | **0.12** | 1.45 | -3.2 | 3.5 | 0.938 | **-3.55** | 0.38 | -4.4 | -2.7 | <0.001 |
| Time | **-0.39** | 0.54 | -1.6 | 0.9 | 0.493 | **-0.77** | 1.27 | -3.8 | 2.2 | 0.561 |
| **Life stage** (Stage 1: unmarried as referent) |  |  |  |  |  |  |  |  |  |  |
| Stage 2: Married under 3 years |  |  |  |  |  | 2.31 | 1.79 | -1.9 | 6.6 | 0.237 |
| Stage 3: Married 3 years or more, age 49 and younger |  |  |  |  |  | -1.74 | 1.24 | -4.7 | 1.2 | 0.203 |
| Stage 4: Over 49 years, any marital status |  |  |  |  |  | -1.47 | 1.52 | -5.1 | 2.1 | 0.365 |
| **Education completed** (primary or less as referent) |  |  |  |  |  |  |  |  |  |  |
| Secondary |  |  |  |  |  | 1.25 | 0.85 | -0.8 | 3.3 | 0.186 |
| Post-secondary |  |  |  |  |  | 0.42 | 1.16 | -2.3 | 3.2 | 0.726 |
| **Self-rated physical health** (poor health as referent) |  |  |  |  |  |  |  |  |  |  |
| Fair |  |  |  |  |  | 0.24 | 0.91 | -1.9 | 2.4 | 0.803 |
| Good |  |  |  |  |  | 0.35 | 0.93 | -1.8 | 2.5 | 0.714 |
| Very good |  |  |  |  |  | -0.96 | 1.09 | -3.5 | 1.6 | 0.412 |
| Excellent |  |  |  |  |  | 0.89 | 0.83 | -1.1 | 2.9 | 0.317 |
| **Unshared latrine** |  |  |  |  |  | 1.19 | 1.53 | -2.4 | 4.8 | 0.462 |
| **Latrine is lockable** |  |  |  |  |  | 2.03 | 2.14 | -3.0 | 7.1 | 0.374 |
| **Sufficient latrine lighting inside latrine** |  |  |  |  |  | 1.25 | 1.01 | -1.1 | 3.6 | 0.256 |
| **Sufficient latrine lighting along way to latrine** |  |  |  |  |  | -2.31 | 0.78 | -4.2 | -0.5 | 0.021 |
| **Physically challenging to access or use latrine** |  |  |  |  |  | -2.43 | 2.36 | -8.0 | 3.2 | 0.338 |
| Models are clustered by neighborhood (n = 8) | | | | | | | | | | |

**Sanitation-related empowerment resources are associated with women’s well-being, anxiety, and depression: findings from Bangladesh, India, Senegal, and Uganda**

Thea Mink, Madeleine Patrick, Amelia Conrad, Tanvir Ahmed, Srishty Arun, Vinod Ramanarayanan, Niladri Chakraborti, Y. Malini Reddy, Abhilaasha Nagarajan, Tanushree Bhan, Sheela S. Sinharoy, Bethany A. Caruso

| **Table S6.** Association between sanitation-related empowerment resources sub-domains and **well-being (WHO-5)**, in the Kampala sample (n = 509) | | | | | | | | | | |
| --- | --- | --- | --- | --- | --- | --- | --- | --- | --- | --- |
|  | Sanitation-related empowerment resources | | | | | Sanitation-related empowerment resources +  individual characteristics +  sanitation environment +  city | | | | |
|  | **Model 1 \| Unadjusted** | | | | | **Model 3 \| Adjusted** | | | | |
| **Parameter** | Estimate | SE | (95% CI) | | p value | Estimate | SE | (95% CI) | | p value |
| **Intercept** | 5.47 | 3.55 | -2.1 | 13.1 | 0.145 | -0.86 | 3.43 | -8.2 | 6.5 | 0.805 |
| **Sanitation-related empowerment sub-domains** |  |  |  |  |  |  |  |  |  |  |
| Bodily Integrity | **1.68** | 0.65 | 0.3 | 3.1 | 0.021 | **0.62** | 0.71 | -0.9 | 2.1 | 0.399 |
| Safety and Security | **0.32** | 0.79 | -1.4 | 2.0 | 0.689 | **0.99** | 0.64 | -0.4 | 2.4 | 0.146 |
| Privacy | **2.52** | 0.61 | 1.2 | 3.8 | 0.001 | **2.69** | 0.59 | 1.4 | 4.0 | <0.001 |
| Time | **-1.07** | 0.43 | -2.0 | -0.1 | 0.027 | **-0.90** | 0.45 | -1.9 | 0.1 | 0.062 |
| **Life stage** (Stage 1: Unmarried, age 49 and younger as referent) |  |  |  |  |  |  |  |  |  |  |
| Stage 2: Married under 3 years |  |  |  |  |  | 1.03 | 1.47 | -2.1 | 4.2 | 0.494 |
| Stage 3: Married 3 years or more, age 49 and younger |  |  |  |  |  | -0.74 | 0.55 | -1.9 | 0.4 | 0.204 |
| Stage 4: Over 49 years, any marital status |  |  |  |  |  | -1.78 | 1.11 | -4.2 | 0.6 | 0.133 |
| **Education completed** (primary or less as referent) |  |  |  |  |  |  |  |  |  |  |
| Secondary |  |  |  |  |  | 0.63 | 0.43 | -0.3 | 1.6 | 0.164 |
| Post-secondary |  |  |  |  |  | 1.45 | 0.53 | 0.3 | 2.6 | 0.016 |
| **Self-rated physical health** (poor health as referent) |  |  |  |  |  |  |  |  |  |  |
| Fair |  |  |  |  |  | 3.23 | 1.11 | 0.9 | 5.6 | 0.011 |
| Good |  |  |  |  |  | 5.42 | 0.96 | 3.4 | 7.5 | <0.001 |
| Very good |  |  |  |  |  | 8.23 | 1.22 | 5.6 | 10.8 | <0.001 |
| Excellent |  |  |  |  |  | 8.50 | 1.37 | 5.6 | 11.4 | <0.001 |
| **Unshared latrine** |  |  |  |  |  | 0.59 | 0.34 | -0.1 | 1.3 | 0.104 |
| **Latrine is lockable** |  |  |  |  |  | -1.23 | 0.47 | -2.2 | -0.2 | 0.019 |
| **Sufficient latrine lighting inside latrine** |  |  |  |  |  | 0.78 | 0.59 | -0.5 | 2.0 | 0.205 |
| **Sufficient latrine lighting along way to latrine** |  |  |  |  |  | 0.41 | 0.76 | -1.2 | 2.0 | 0.596 |
| **Physically challenging to access or use latrine** |  |  |  |  |  | 0.95 | 0.65 | -0.4 | 2.4 | 0.167 |
| Models are clustered by neighborhood (n = 15) |  |  |  |  |  |  |  |  |  |  |

**Sanitation-related empowerment resources are associated with women’s well-being, anxiety, and depression: findings from Bangladesh, India, Senegal, and Uganda**

Thea Mink, Madeleine Patrick, Amelia Conrad, Tanvir Ahmed, Srishty Arun, Vinod Ramanarayanan, Niladri Chakraborti, Y. Malini Reddy, Abhilaasha Nagarajan, Tanushree Bhan, Sheela S. Sinharoy, Bethany A. Caruso

| **Table S7.** Association between sanitation-related empowerment resources sub-domains and **depression (PHQ-2)**, in the Meherpur sample (n = 390) | | | | | | | | | | |
| --- | --- | --- | --- | --- | --- | --- | --- | --- | --- | --- |
|  | Sanitation-related empowerment resources | | | | | Sanitation-related empowerment resources +  individual characteristics +  sanitation environment +  city | | | | |
|  | **Model 1 \| Unadjusted** | | | | | **Model 3 \| Adjusted** | | | | |
| **Parameter** | Estimate | SE | (95% CI) | | p value | Estimate | SE | (95% CI) | | p value |
| **Intercept** | 5.15 | 1.10 | 2.9 | 7.4 | <0.000 | 4.16 | 1.29 | 1.5 | 6.8 | 0.003 |
| **Sanitation-related empowerment sub-domains** |  |  |  |  |  |  |  |  |  |  |
| Bodily Integrity | **-0.66** | 0.28 | -1.2 | -0.1 | 0.022 | **-0.11** | 0.28 | -0.7 | 0.5 | 0.692 |
| Safety and Security | **-0.24** | 0.27 | -0.8 | 0.3 | 0.376 | **-0.05** | 0.25 | -0.6 | 0.5 | 0.853 |
| Privacy | **-0.04** | 0.15 | -0.4 | 0.3 | 0.787 | **-0.13** | 0.11 | -0.4 | 0.1 | 0.219 |
| Time | **-0.06** | 0.18 | -0.4 | 0.3 | 0.753 | **0.00** | 0.19 | -0.4 | 0.4 | 0.984 |
| **Life stage** (Stage 1: Unmarried, age 49 and younger and referent) | |  |  |  |  |  |  |  |  |  |
| Stage 2: Married under 3 years |  |  |  |  |  | -0.13 | 0.61 | -1.4 | 1.1 | 0.833 |
| Stage 3: Married 3 years or more, age 49 and younger |  |  |  |  |  | 0.10 | 0.44 | -0.8 | 1.0 | 0.829 |
| Stage 4: Over 49 years, any marital status |  |  |  |  |  | 0.52 | 0.52 | -0.5 | 1.6 | 0.320 |
| **Education completed** (primary or less as referent) |  |  |  |  |  |  |  |  |  |  |
| Secondary |  |  |  |  |  | -0.42 | 0.17 | -0.8 | -0.1 | 0.019 |
| Post-secondary |  |  |  |  |  | -0.52 | 0.23 | -1.0 | -0.1 | 0.030 |
| **Self-rated physical health** (poor health as referent) |  |  |  |  |  |  |  |  |  |  |
| Fair |  |  |  |  |  | -0.14 | 0.67 | -1.5 | 1.2 | 0.830 |
| Good |  |  |  |  |  | -0.77 | 0.72 | -2.2 | 0.7 | 0.289 |
| Very good |  |  |  |  |  | -1.62 | 0.71 | -3.1 | -0.2 | 0.028 |
| Excellent |  |  |  |  |  | -1.54 | 0.70 | -3.0 | -0.1 | 0.035 |
| **City** |  |  |  |  |  |  |  |  |  |  |
| **Unshared latrine** |  |  |  |  |  | 0.00 | 0.22 | -0.4 | 0.5 | 0.993 |
| **Latrine is lockable** |  |  |  |  |  | 0.36 | 0.24 | -0.1 | 0.8 | 0.148 |
| **Sufficient latrine lighting inside latrine** |  |  |  |  |  | 0.13 | 0.30 | -0.5 | 0.7 | 0.671 |
| **Sufficient latrine lighting along way to latrine** |  |  |  |  |  | -1.31 | 0.52 | -2.4 | -0.3 | 0.017 |
| **Physically challenging to access or use latrine** |  |  |  |  |  | 0.42 | 0.29 | -0.2 | 1.0 | 0.162 |
| Models are clustered by neighborhood (n = 34) |  |  |  |  |  |  |  |  |  |  |

**Sanitation-related empowerment resources are associated with women’s well-being, anxiety, and depression: findings from Bangladesh, India, Senegal, and Uganda**

Thea Mink, Madeleine Patrick, Amelia Conrad, Tanvir Ahmed, Srishty Arun, Vinod Ramanarayanan, Niladri Chakraborti, Y. Malini Reddy, Abhilaasha Nagarajan, Tanushree Bhan, Sheela S. Sinharoy, Bethany A. Caruso

| **Table S8.** Association between sanitation-related empowerment resources sub-domains and **depression (PHQ-2)**, in the Saidpur sample (n = 659) | | | | | | | | | | |
| --- | --- | --- | --- | --- | --- | --- | --- | --- | --- | --- |
|  | Sanitation-related empowerment resources | | | | | Sanitation-related empowerment resources +  individual characteristics +  sanitation environment +  city | | | | |
|  | **Model 1 \| Unadjusted** | | | | | **Model 3 \| Adjusted** | | | | |
| **Parameter** | Estimate | SE | (95% CI) | | p value | Estimate | SE | (95% CI) | | p value |
| **Intercept** | 6.32 | 1.01 | 4.3 | 8.3 | <0.000 | 9.17 | 1.41 | 6.3 | 12.0 | <0.001 |
| **Sanitation-related empowerment sub-domains** |  |  |  |  |  |  |  |  |  |  |
| Bodily Integrity | **-0.76** | 0.25 | -1.3 | -0.3 | 0.004 | **-1.07** | 0.27 | -1.6 | -0.5 | <0.001 |
| Safety and Security | **-0.27** | 0.21 | -0.7 | 0.2 | 0.209 | **-0.36** | 0.22 | -0.8 | 0.1 | 0.104 |
| Privacy | **-0.06** | 0.21 | -0.5 | 0.4 | 0.771 | **-0.44** | 0.26 | -1.0 | 0.1 | 0.101 |
| Time | **-0.47** | 0.11 | -0.7 | -0.3 | <0.000 | **-0.38** | 0.12 | -0.6 | -0.1 | 0.002 |
| **Life stage** (Stage 1: Unmarried, age 49 and younger as referent) | |  |  |  |  |  |  |  |  |  |
| Stage 2: Married under 3 years |  |  |  |  |  | -0.02 | 0.21 | -0.4 | 0.4 | 0.929 |
| Stage 3: Married 3 years or more, age 49 and younger |  |  |  |  |  | -0.03 | 0.12 | -0.3 | 0.2 | 0.785 |
| Stage 4: Over 49 years, any marital status |  |  |  |  |  | -0.21 | 0.18 | -0.6 | 0.2 | 0.254 |
| **Education completed** (primary or less as referent) |  |  |  |  |  |  |  |  |  |  |
| Secondary |  |  |  |  |  | 0.04 | 0.09 | -0.1 | 0.2 | 0.682 |
| Post-secondary |  |  |  |  |  | 0.06 | 0.14 | -0.2 | 0.3 | 0.640 |
| **Self-rated physical health** (poor health as referent) |  |  |  |  |  |  |  |  |  |  |
| Fair |  |  |  |  |  | -0.69 | 0.37 | -1.4 | 0.1 | 0.070 |
| Good |  |  |  |  |  | -1.02 | 0.39 | -1.8 | -0.2 | 0.012 |
| Very good |  |  |  |  |  | -1.08 | 0.40 | -1.9 | -0.3 | 0.009 |
| Excellent |  |  |  |  |  | -1.05 | 0.40 | -1.9 | -0.2 | 0.012 |
| **City** |  |  |  |  |  |  |  |  |  |  |
| **Unshared latrine** |  |  |  |  |  | 0.36 | 0.15 | 0.1 | 0.7 | 0.019 |
| **Latrine is lockable** |  |  |  |  |  | 0.15 | 0.22 | -0.3 | 0.6 | 0.482 |
| **Sufficient latrine lighting inside latrine** |  |  |  |  |  | 0.06 | 0.22 | -0.4 | 0.5 | 0.777 |
| **Sufficient latrine lighting along way to latrine** |  |  |  |  |  | 0.29 | 0.31 | -0.3 | 0.9 | 0.346 |
| **Physically challenging to access or use latrine** |  |  |  |  |  | -0.49 | 0.46 | -1.4 | 0.4 | 0.293 |
| Models are clustered by neighborhood (n = 43) |  |  |  |  |  |  |  |  |  |  |

**Sanitation-related empowerment resources are associated with women’s well-being, anxiety, and depression: findings from Bangladesh, India, Senegal, and Uganda**

Thea Mink, Madeleine Patrick, Amelia Conrad, Tanvir Ahmed, Srishty Arun, Vinod Ramanarayanan, Niladri Chakraborti, Y. Malini Reddy, Abhilaasha Nagarajan, Tanushree Bhan, Sheela S. Sinharoy, Bethany A. Caruso

| **Table S9.** Association between sanitation-related empowerment resources sub-domains and **depression (PHQ-2)**, in the Tiruchirappalli sample (n = 304) | | | | | | | | | | |
| --- | --- | --- | --- | --- | --- | --- | --- | --- | --- | --- |
|  | Sanitation-related empowerment resources | | | | | Sanitation-related empowerment resources +  individual characteristics +  sanitation environment +  city | | | | |
|  | **Model 1 \| Unadjusted** | | | | | **Model 3 \| Adjusted** | | | | |
| **Parameter** | Estimate | SE | (95% CI) | | p value | Estimate | SE | (95% CI) | | p value |
| **Intercept** | 3.39 | 0.53 | 2.2 | 4.5 | <0.000 | 4.03 | 0.82 | 2.2 | 5.8 | <0.001 |
| **Sanitation-related empowerment sub-domains** |  |  |  |  |  |  |  |  |  |  |
| Bodily Integrity | -0.41 | 0.33 | -1.1 | 0.3 | 0.235 | **-0.26** | 0.35 | -1.0 | 0.5 | 0.478 |
| Safety and Security | 0.86 | 0.24 | 0.3 | 1.4 | 0.004 | **0.61** | 0.32 | -0.1 | 1.3 | 0.078 |
| Privacy | -0.13 | 0.34 | -0.9 | 0.6 | 0.712 | **-0.21** | 0.30 | -0.9 | 0.5 | 0.500 |
| Time | -0.65 | 0.22 | -1.1 | -0.2 | 0.013 | **-0.39** | 0.25 | -0.9 | 0.1 | 0.138 |
| **Life stage** (Stage 1: Unmarried, age 49 and younger as referent) | |  |  |  |  |  |  |  |  |  |
| Stage 2: Married under 3 years |  |  |  |  |  | 0.01 | 0.40 | -0.9 | 0.9 | 0.974 |
| Stage 3: Married 3 years or more, age 49 and younger |  |  |  |  |  | -0.08 | 0.13 | -0.4 | 0.2 | 0.553 |
| Stage 4: Over 49 years, any marital status |  |  |  |  |  | -0.52 | 0.95 | -2.6 | 1.5 | 0.593 |
| **Education completed** (primary or less as referent) |  |  |  |  |  |  |  |  |  |  |
| Secondary |  |  |  |  |  | -0.06 | 0.09 | -0.3 | 0.1 | 0.502 |
| Post-secondary |  |  |  |  |  | -0.14 | 0.12 | -0.4 | 0.1 | 0.276 |
| **Self-rated physical health** (poor health as referent) |  |  |  |  |  |  |  |  |  |  |
| Fair |  |  |  |  |  | -0.33 | 0.52 | -1.5 | 0.8 | 0.542 |
| Good |  |  |  |  |  | -0.38 | 0.53 | -1.5 | 0.8 | 0.487 |
| Very good |  |  |  |  |  | -0.18 | 0.61 | -1.5 | 1.1 | 0.771 |
| Excellent |  |  |  |  |  | -0.22 | 0.59 | -1.5 | 1.1 | 0.723 |
| **City** |  |  |  |  |  |  |  |  |  |  |
| **Unshared latrine** |  |  |  |  |  | 0.07 | 0.23 | -0.4 | 0.6 | 0.776 |
| **Latrine is lockable** |  |  |  |  |  | -0.08 | 0.22 | -0.6 | 0.4 | 0.744 |
| **Sufficient latrine lighting inside latrine** |  |  |  |  |  | 0.60 | 0.45 | -0.4 | 1.6 | 0.207 |
| **Sufficient latrine lighting along way to latrine** |  |  |  |  |  | -1.27 | 0.53 | -2.4 | -0.1 | 0.033 |
| **Physically challenging to access or use latrine** |  |  |  |  |  | 0.92 | 0.17 | 0.6 | 1.3 | <0.001 |
| Models are clustered by neighborhood (n = 13) |  |  |  |  |  |  |  |  |  |  |

**Sanitation-related empowerment resources are associated with women’s well-being, anxiety, and depression: findings from Bangladesh, India, Senegal, and Uganda**

Thea Mink, Madeleine Patrick, Amelia Conrad, Tanvir Ahmed, Srishty Arun, Vinod Ramanarayanan, Niladri Chakraborti, Y. Malini Reddy, Abhilaasha Nagarajan, Tanushree Bhan, Sheela S. Sinharoy, Bethany A. Caruso

| **Table S10.** Association between sanitation-related empowerment resources sub-domains and **depression (PHQ-2)**, in the Dakar sample (n = 260) | | | | | | | | | | |
| --- | --- | --- | --- | --- | --- | --- | --- | --- | --- | --- |
|  | Sanitation-related empowerment resources | | | | | Sanitation-related empowerment resources +  individual characteristics +  sanitation environment +  city | | | | |
|  | **Model 1 \| Unadjusted** | | | | | **Model 3 \| Adjusted** | | | | |
| **Parameter** | Estimate | SE | (95% CI) | | p value | Estimate | SE | (95% CI) | | p value |
| **Intercept** | 6.23 | 0.74 | 4.5 | 7.9 | <0.000 | 5.09 | 0.69 | 3.5 | 6.7 | <0.001 |
| **Sanitation-related empowerment sub-domains** |  |  |  |  |  |  |  |  |  |  |
| Bodily Integrity | **-0.22** | 0.09 | -0.4 | 0.0 | 0.035 | **-0.33** | 0.08 | -0.5 | -0.1 | 0.005 |
| Safety and Security | **-0.20** | 0.20 | -0.6 | 0.3 | 0.349 | **0.02** | 0.25 | -0.6 | 0.6 | 0.924 |
| Privacy | **-1.22** | 0.16 | -1.6 | -0.9 | <0.000 | **-0.87** | 0.20 | -1.3 | -0.4 | 0.003 |
| Time | **0.14** | 0.10 | -0.1 | 0.4 | 0.231 | **0.08** | 0.11 | -0.2 | 0.3 | 0.500 |
| **Life stage** (Stage 1: Unmarried, age 49 and younger as referent) | |  |  |  |  |  |  |  |  |  |
| Stage 2: Married under 3 years |  |  |  |  |  | -0.24 | 0.13 | -0.6 | 0.1 | 0.110 |
| Stage 3: Married 3 years or more, age 49 and younger |  |  |  |  |  | 0.06 | 0.04 | 0.0 | 0.1 | 0.187 |
| Stage 4: Over 49 years, any marital status |  |  |  |  |  | -0.14 | 0.10 | -0.4 | 0.1 | 0.178 |
| **Education completed** (primary or less as referent) |  |  |  |  |  |  |  |  |  |  |
| Secondary |  |  |  |  |  | -0.10 | 0.05 | -0.2 | 0.0 | 0.068 |
| Post-secondary |  |  |  |  |  | -0.16 | 0.09 | -0.4 | 0.0 | 0.113 |
| **Self-rated physical health** (poor health as referent) |  |  |  |  |  |  |  |  |  |  |
| Fair |  |  |  |  |  | -0.68 | 0.32 | -1.4 | 0.1 | 0.068 |
| Good |  |  |  |  |  | -0.72 | 0.33 | -1.5 | 0.1 | 0.064 |
| Very good |  |  |  |  |  | -0.72 | 0.39 | -1.6 | 0.2 | 0.107 |
| Excellent |  |  |  |  |  | -0.81 | 0.28 | -1.5 | -0.2 | 0.022 |
| **City** |  |  |  |  |  |  |  |  |  |  |
| **Unshared latrine** |  |  |  |  |  | -0.10 | 0.14 | -0.4 | 0.2 | 0.509 |
| **Latrine is lockable** |  |  |  |  |  | 0.65 | 0.24 | 0.1 | 1.2 | 0.032 |
| **Sufficient latrine lighting inside latrine** |  |  |  |  |  | -0.28 | 0.21 | -0.8 | 0.2 | 0.220 |
| **Sufficient latrine lighting along way to latrine** |  |  |  |  |  | 0.05 | 0.12 | -0.2 | 0.3 | 0.690 |
| **Physically challenging to access or use latrine** |  |  |  |  |  | -0.10 | 0.19 | -0.5 | 0.3 | 0.623 |
| Models are clustered by neighborhood (n = 8) |  |  |  |  |  |  |  |  |  |  |

**Sanitation-related empowerment resources are associated with women’s well-being, anxiety, and depression: findings from Bangladesh, India, Senegal, and Uganda**

Thea Mink, Madeleine Patrick, Amelia Conrad, Tanvir Ahmed, Srishty Arun, Vinod Ramanarayanan, Niladri Chakraborti, Y. Malini Reddy, Abhilaasha Nagarajan, Tanushree Bhan, Sheela S. Sinharoy, Bethany A. Caruso

| **Table S11.** Association between sanitation-related empowerment resources sub-domains and **depression (PHQ-2)**, in the Kampala sample (n = 509) | | | | | | | | | | |
| --- | --- | --- | --- | --- | --- | --- | --- | --- | --- | --- |
|  | Sanitation-related empowerment resources | | | | | Sanitation-related empowerment resources +  individual characteristics +  sanitation environment +  city | | | | |
|  | **Model 1 \| Unadjusted** | | | | | **Model 3 \| Adjusted** | | | | |
| **Parameter** | Estimate | SE | (95% CI) | | p value | Estimate | SE | (95% CI) | | p value |
| **Intercept** | 5.53 | 0.50 | 4.4 | 6.6 | <0.000 | 6.75 | 0.53 | 5.6 | 7.9 | <0.001 |
| **Sanitation-related empowerment sub-domains** |  |  |  |  |  |  |  |  |  |  |
| Bodily Integrity | **-0.30** | 0.14 | -0.6 | 0.0 | 0.058 | **-0.22** | 0.11 | -0.4 | 0.0 | 0.063 |
| Safety and Security | **-0.07** | 0.18 | -0.4 | 0.3 | 0.715 | **-0.25** | 0.14 | -0.6 | 0.1 | 0.105 |
| Privacy | **-0.73** | 0.15 | -1.1 | -0.4 | <0.000 | **-0.82** | 0.19 | -1.2 | -0.4 | 0.001 |
| Time | **-0.07** | 0.12 | -0.3 | 0.2 | 0.587 | **0.06** | 0.12 | -0.2 | 0.3 | 0.641 |
| **Life stage** (Stage 1: Unmarried, age 49 and younger as referent) | |  |  |  |  |  |  |  |  |  |
| Stage 2: Married under 3 years |  |  |  |  |  | -0.56 | 0.28 | -1.2 | 0.0 | 0.062 |
| Stage 3: Married 3 years or more, age 49 and younger |  |  |  |  |  | 0.04 | 0.13 | -0.2 | 0.3 | 0.784 |
| Stage 4: Over 49 years, any marital status |  |  |  |  |  | -0.08 | 0.19 | -0.5 | 0.3 | 0.667 |
| **Education completed** (primary or less as referent) |  |  |  |  |  |  |  |  |  |  |
| Secondary |  |  |  |  |  | 0.09 | 0.16 | -0.3 | 0.4 | 0.593 |
| Post-secondary |  |  |  |  |  | -0.11 | 0.19 | -0.5 | 0.3 | 0.566 |
| **Self-rated physical health** (poor health as referent) |  |  |  |  |  |  |  |  |  |  |
| Fair |  |  |  |  |  | -0.28 | 0.39 | -1.1 | 0.6 | 0.482 |
| Good |  |  |  |  |  | -1.04 | 0.28 | -1.6 | -0.4 | 0.002 |
| Very good |  |  |  |  |  | -1.74 | 0.33 | -2.4 | -1.0 | <0.001 |
| Excellent |  |  |  |  |  | -1.51 | 0.29 | -2.1 | -0.9 | <0.001 |
| **City** |  |  |  |  |  |  |  |  |  |  |
|  |  |  |  |  |  | -0.26 | 0.12 | -0.5 | 0.0 | 0.050 |
| **Unshared latrine** |  |  |  |  |  | 0.15 | 0.19 | -0.3 | 0.6 | 0.456 |
| **Latrine is lockable** |  |  |  |  |  | 0.30 | 0.21 | -0.1 | 0.7 | 0.174 |
| **Sufficient latrine lighting inside latrine** |  |  |  |  |  | 0.10 | 0.22 | -0.4 | 0.6 | 0.664 |
| **Sufficient latrine lighting along way to latrine** |  |  |  |  |  | -0.38 | 0.18 | -0.8 | 0.0 | 0.054 |
| **Physically challenging to access or use latrine** |  |  |  |  |  |  |  |  |  |  |
| Models are clustered by neighborhood (n = 15) |  |  |  |  |  |  |  |  |  |  |

**Sanitation-related empowerment resources are associated with women’s well-being, anxiety, and depression: findings from Bangladesh, India, Senegal, and Uganda**

Thea Mink, Madeleine Patrick, Amelia Conrad, Tanvir Ahmed, Srishty Arun, Vinod Ramanarayanan, Niladri Chakraborti, Y. Malini Reddy, Abhilaasha Nagarajan, Tanushree Bhan, Sheela S. Sinharoy, Bethany A. Caruso

| **Table S12.** Association between sanitation-related empowerment resources sub-domains and **anxiety (GAD-2)**, in the Meherpur sample (n = 390) | | | | | | | | | | |
| --- | --- | --- | --- | --- | --- | --- | --- | --- | --- | --- |
|  | Sanitation-related empowerment resources | | | | | Sanitation-related empowerment resources +  individual characteristics +  sanitation environment +  city | | | | |
|  | **Model 1 \| Unadjusted** | | | | | **Model 3 \| Adjusted** | | | | |
| **Parameter** | Estimate | SE | (95% CI) | | p value | Estimate | SE | (95% CI) | | p value |
| **Intercept** | 5.13 | 1.02 | 3.0 | 7.2 | 0 | 6.59 | 1.34 | 3.9 | 9.3 | <0.001 |
| **Sanitation-related empowerment sub-domains** |  |  |  |  |  |  |  |  |  |  |
| Bodily Integrity | **-0.65** | 0.23 | -1.1 | -0.2 | 0.008 | **-0.25** | 0.26 | -0.8 | 0.3 | 0.346 |
| Safety and Security | **-0.08** | 0.26 | -0.6 | 0.4 | 0.764 | **-0.14** | 0.25 | -0.6 | 0.4 | 0.561 |
| Privacy | **-0.04** | 0.18 | -0.4 | 0.3 | 0.814 | **-0.11** | 0.12 | -0.4 | 0.1 | 0.374 |
| Time | **-0.24** | 0.16 | -0.6 | 0.1 | 0.143 | **-0.19** | 0.18 | -0.6 | 0.2 | 0.296 |
| **Life stage** (Stage 1: Unmarried, age 49 and younger as referent) | |  |  |  |  |  |  |  |  |  |
| Stage 2: Married under 3 years |  |  |  |  |  | -0.74 | 0.39 | -1.5 | 0.1 | 0.069 |
| Stage 3: Married 3 years or more, age 49 and younger |  |  |  |  |  | -0.62 | 0.29 | -1.2 | 0.0 | 0.038 |
| Stage 4: Over 49 years, any marital status |  |  |  |  |  | 0.00 | 0.42 | -0.9 | 0.9 | 0.991 |
| **Education completed** (primary or less as referent) |  |  |  |  |  |  |  |  |  |  |
| Secondary |  |  |  |  |  | -0.31 | 0.24 | -0.8 | 0.2 | 0.201 |
| Post-secondary |  |  |  |  |  | -0.45 | 0.29 | -1.0 | 0.1 | 0.132 |
| **Self-rated physical health** (poor health as referent) |  |  |  |  |  |  |  |  |  |  |
| Fair |  |  |  |  |  | -0.53 | 0.75 | -2.0 | 1.0 | 0.485 |
| Good |  |  |  |  |  | -1.19 | 0.80 | -2.8 | 0.4 | 0.148 |
| Very good |  |  |  |  |  | -1.75 | 0.81 | -3.4 | -0.1 | 0.038 |
| Excellent |  |  |  |  |  | -1.80 | 0.86 | -3.6 | 0.0 | 0.045 |
| **Social Support** |  |  |  |  |  | -0.42 | 0.15 | -0.7 | -0.1 | 0.007 |
| **Unshared latrine** |  |  |  |  |  | 0.00 | 0.20 | -0.4 | 0.4 | 0.991 |
| **Latrine is lockable** |  |  |  |  |  | 0.71 | 0.20 | 0.3 | 1.1 | 0.001 |
| **Sufficient latrine lighting inside latrine** |  |  |  |  |  | 0.35 | 0.34 | -0.3 | 1.0 | 0.311 |
| **Sufficient latrine lighting along way to latrine** |  |  |  |  |  | -0.59 | 0.36 | -1.3 | 0.1 | 0.111 |
| **Physically challenging to access or use latrine** |  |  |  |  |  | 0.35 | 0.29 | -0.2 | 0.9 | 0.236 |
| Models are clustered by neighborhood (n = 34) |  |  |  |  |  |  |  |  |  |  |

**Sanitation-related empowerment resources are associated with women’s well-being, anxiety, and depression: findings from Bangladesh, India, Senegal, and Uganda**

Thea Mink, Madeleine Patrick, Amelia Conrad, Tanvir Ahmed, Srishty Arun, Vinod Ramanarayanan, Niladri Chakraborti, Y. Malini Reddy, Abhilaasha Nagarajan, Tanushree Bhan, Sheela S. Sinharoy, Bethany A. Caruso

| **Table S13.** Association between sanitation-related empowerment resources sub-domains and **anxiety (GAD-2)**, in the Saidpur sample (n = 659) | | | | | | | | | | |
| --- | --- | --- | --- | --- | --- | --- | --- | --- | --- | --- |
|  | Sanitation-related empowerment resources | | | | | Sanitation-related empowerment resources +  individual characteristics +  sanitation environment +  city | | | | |
|  | **Model 1 \| Unadjusted** | | | | | **Model 3 \| Adjusted** | | | | |
| **Parameter** | Estimate | SE | (95% CI) | | p value | Estimate | SE | (95% CI) | | p value |
| **Intercept** | 5.78 | 1.04 | 3.7 | 7.9 | <0.000 | 7.43 | 1.45 | 4.5 | 10.4 | <0.001 |
| **Sanitation-related empowerment sub-domains** |  |  |  |  |  |  |  |  |  |  |
| Bodily Integrity | **-0.80** | 0.26 | -1.3 | -0.3 | 0.004 | **-0.93** | 0.23 | -1.4 | -0.5 | <0.001 |
| Safety and Security | **0.10** | 0.24 | -0.4 | 0.6 | 0.670 | **0.07** | 0.24 | -0.4 | 0.6 | 0.769 |
| Privacy | **-0.19** | 0.27 | -0.7 | 0.4 | 0.492 | **-0.43** | 0.31 | -1.1 | 0.2 | 0.175 |
| Time | **-0.55** | 0.13 | -0.8 | -0.3 | <0.000 | **-0.48** | 0.13 | -0.8 | -0.2 | 0.001 |
| **Life stage** (Stage 1: Unmarried, age 49 and younger as referent) | |  |  |  |  |  |  |  |  |  |
| Stage 2: Married under 3 years |  |  |  |  |  | 0.14 | 0.21 | -0.3 | 0.6 | 0.489 |
| Stage 3: Married 3 years or more, age 49 and younger |  |  |  |  |  | 0.13 | 0.12 | -0.1 | 0.4 | 0.297 |
| Stage 4: Over 49 years, any marital status |  |  |  |  |  | 0.02 | 0.19 | -0.4 | 0.4 | 0.903 |
| **Education completed** (primary or less as referent) |  |  |  |  |  |  |  |  |  |  |
| Secondary |  |  |  |  |  | 0.10 | 0.08 | -0.1 | 0.3 | 0.230 |
| Post-secondary |  |  |  |  |  | 0.11 | 0.15 | -0.2 | 0.4 | 0.446 |
| **Self-rated physical health** (poor health as referent) |  |  |  |  |  |  |  |  |  |  |
| Fair |  |  |  |  |  | -0.95 | 0.68 | -2.3 | 0.4 | 0.173 |
| Good |  |  |  |  |  | -1.22 | 0.68 | -2.6 | 0.2 | 0.082 |
| Very good |  |  |  |  |  | -1.29 | 0.69 | -2.7 | 0.1 | 0.066 |
| Excellent |  |  |  |  |  | -1.14 | 0.62 | -2.4 | 0.1 | 0.075 |
| **Social Support** |  |  |  |  |  | 0.06 | 0.06 | -0.1 | 0.2 | 0.375 |
| **Unshared latrine** |  |  |  |  |  | 0.22 | 0.15 | -0.1 | 0.5 | 0.139 |
| **Latrine is lockable** |  |  |  |  |  | -0.02 | 0.28 | -0.6 | 0.5 | 0.951 |
| **Sufficient latrine lighting inside latrine** |  |  |  |  |  | 0.04 | 0.20 | -0.4 | 0.4 | 0.825 |
| **Sufficient latrine lighting along way to latrine** |  |  |  |  |  | 0.27 | 0.27 | -0.3 | 0.8 | 0.317 |
| **Physically challenging to access or use latrine** |  |  |  |  |  | -0.57 | 0.42 | -1.4 | 0.3 | 0.179 |
| Models are clustered by neighborhood (n = 43) |  |  |  |  |  |  |  |  |  |  |

**Sanitation-related empowerment resources are associated with women’s well-being, anxiety, and depression: findings from Bangladesh, India, Senegal, and Uganda**

Thea Mink, Madeleine Patrick, Amelia Conrad, Tanvir Ahmed, Srishty Arun, Vinod Ramanarayanan, Niladri Chakraborti, Y. Malini Reddy, Abhilaasha Nagarajan, Tanushree Bhan, Sheela S. Sinharoy, Bethany A. Caruso

| **Table S14.** Association between sanitation-related empowerment resources sub-domains and **anxiety (GAD-2)**, in the Tiruchirappalli sample (n = 304) | | | | | | | | | | |
| --- | --- | --- | --- | --- | --- | --- | --- | --- | --- | --- |
|  | Sanitation-related empowerment resources | | | | | Sanitation-related empowerment resources +  individual characteristics +  sanitation environment +  city | | | | |
|  | **Model 1 \| Unadjusted** | | | | | **Model 3 \| Adjusted** | | | | |
| **Parameter** | Estimate | SE | (95% CI) | | p value | Estimate | SE | (95% CI) | | p value |
| **Intercept** | 7.75 | 0.78 | 6.1 | 9.4 | <0.000 | 8.15 | 2.13 | 3.5 | 12.8 | 0.002 |
| **Sanitation-related empowerment sub-domains** |  |  |  |  |  |  |  |  |  |  |
| Bodily Integrity | **-0.84** | 0.24 | -1.4 | -0.3 | 0.005 | **-0.64** | 0.26 | -1.2 | -0.1 | 0.030 |
| Safety and Security | **-0.77** | 0.23 | -1.3 | -0.3 | 0.006 | **-0.74** | 0.17 | -1.1 | -0.4 | 0.001 |
| Privacy | **-0.15** | 0.26 | -0.7 | 0.4 | 0.568 | **-0.25** | 0.26 | -0.8 | 0.3 | 0.359 |
| Time | **-0.07** | 0.15 | -0.4 | 0.2 | 0.650 | **0.16** | 0.19 | -0.2 | 0.6 | 0.400 |
| **Life stage** (Stage 1: Unmarried, age 49 and younger as referent) | |  |  |  |  |  |  |  |  |  |
| Stage 2: Married under 3 years |  |  |  |  |  | -0.03 | 0.52 | -1.2 | 1.1 | 0.957 |
| Stage 3: Married 3 years or more, age 49 and younger |  |  |  |  |  | 0.27 | 0.25 | -0.3 | 0.8 | 0.300 |
| Stage 4: Over 49 years, any marital status |  |  |  |  |  | -0.51 | 0.26 | -1.1 | 0.1 | 0.075 |
| **Education completed** (primary or less as referent) |  |  |  |  |  |  |  |  |  |  |
| Secondary |  |  |  |  |  | 0.13 | 0.14 | -0.2 | 0.4 | 0.398 |
| Post-secondary |  |  |  |  |  | -0.01 | 0.14 | -0.3 | 0.3 | 0.929 |
| **Self-rated physical health** (poor health as referent) |  |  |  |  |  |  |  |  |  |  |
| Fair |  |  |  |  |  | 0.05 | 0.64 | -1.3 | 1.5 | 0.938 |
| Good |  |  |  |  |  | -0.20 | 0.43 | -1.1 | 0.7 | 0.646 |
| Very good |  |  |  |  |  | -0.49 | 0.52 | -1.6 | 0.6 | 0.367 |
| Excellent |  |  |  |  |  | -0.57 | 0.47 | -1.6 | 0.5 | 0.254 |
| **Social Support** |  |  |  |  |  | -0.29 | 0.11 | -0.5 | 0.0 | 0.022 |
| **Unshared latrine** |  |  |  |  |  | 0.07 | 0.19 | -0.3 | 0.5 | 0.703 |
| **Latrine is lockable** |  |  |  |  |  | -0.11 | 0.27 | -0.7 | 0.5 | 0.686 |
| **Sufficient latrine lighting inside latrine** |  |  |  |  |  | 0.66 | 0.29 | 0.0 | 1.3 | 0.042 |
| **Sufficient latrine lighting along way to latrine** |  |  |  |  |  | -1.41 | 1.81 | -5.3 | 2.5 | 0.451 |
| **Physically challenging to access or use latrine** |  |  |  |  |  | 0.72 | 0.18 | 0.3 | 1.1 | 0.002 |
| Models are clustered by neighborhood (n = 13) |  |  |  |  |  |  |  |  |  |  |
